# PheCode-guided multi-modal topic modeling of electronic health records improves disease incidence prediction and GWAS discovery from UK Biobank

**DOI:** 10.1101/2025.05.28.25328511

**Authors:** Ziqi Yang, Ziyang Song, Shadi Zabad, Marc-André Legault, Yue Li

## Abstract

Phenome-wide association studies (PheWAS) rely on disease definitions derived from diagnostic codes, often failing to leverage the full richness of electronic health records (EHR). We present MixEHR-SAGE, a PheCode-guided multi-modal topic model that integrates diagnoses, procedures, and medications to enhance phenotyping from large-scale EHRs. By combining expert-informed priors with probabilistic inference, MixEHR-SAGE identifies over 1,000 interpretable phenotype topics from UK Biobank (UKB) data. Applied to 350,000 individuals with high-quality genetic data, MixEHR-SAGE-derived risk scores accurately predict incident type 2 diabetes (T2D) and leukemia diagnoses. Subsequent genome-wide association studies (GWAS) using these continuous risk scores uncovered novel diasease-associated loci, including *PPP1R15A* for T2D and *JMJD6*/*SRSF2* for leukemia, that were missed by traditional binary case definitions. These results highlight the potential of probabilistic phenotyping from multi-modal EHRs to improve genetic discovery. The MixEHR-SAGE software is publicly available at: https://github.com/li-lab-mcgill/MixEHR-SAGE.

## 1 Introduction

Phenome-wide association studies (PheWAS) are widely used to identify associations between genetic variants and a diverse set of diseases [1, 2]. These studies are enabled by the growing availability of genetic and electronic health record (EHR) data in large cohorts, such as the UK Biobank (UKB). The UKB includes over 500,000 participants with available genotyping data, self-reported pharmacotherapy data and linkage to various health records and registries providing hospitalization diagnostic codes and surgical procedure codes. The extensive phenotypic characterization of UKB participants, in combination to the availability of genetic data, has enabled large-scale PheWAS with results in online repositories (e.g. OpenTargets [3]).

However, medico-administrative records represent an imperfect view of participant’s health, representing a challenge in PheWAS [4]. In PheWAS, researchers commonly rely on diagnostic algorithms that combine International Classification of Diseases (ICD) 9 and 10 codes to define disease status for genetic association testing [5–7]. The most commonly used mapping of ICD codes used in PheWAS are called *Phenotype Codes (PheCodes)* [8]. These PheCodes were defined manually and are organized in categories (e.g. cardiovascular system, digestive, neoplasms, *etc*.), and span most health-related phenotypes. An important limitation of PheCodes is that they only rely on ICD codes and do not leverage complementary sources of information, such as pharmacotherapy data or surgical procedure codes. Furthermore, the validity of PheCodes may vary between studies and datasets due to differences ICD code usage between physicians or institutions, motivating the development of adaptive systems.

Topic models offer a powerful and data-driven approach to create algorithmic disease definitions. They were developed to summarize documents based on a mixture of latent topics defined by statistical word co-occurrences in the field of natural language processing [9, 10]. By analogy, EHRs can be viewed as “documents”, where co-occurring diagnostic codes capture the unobserved disease phenotypes, and patient-level topic mixtures can be interpreted as continuous disease scores. MixEHR uses topic models to infer a joint distribution across various EHR data types – such as diagnostic codes, self-reported pharmacotherapy data, and surgical procedure codes – to more comprehensively capture disease status from complex EHR data [11]. Usually, the use of topic models is hampered by the need for the *post hoc* interpretation of latent disease topics, as they are learned without disease labels. This characteristic is shared with other latent variable models, such as principal component analysis and non-negative matrix factorization which are frequently used to analyze the high-dimensional EHR data and uncover hidden disease representations in the context of phenotyping and risk stratification [12]. Recent advances in topic models developed for EHR data alleviate this problem [13, 14]. Namely, MixEHR-G incorporates expert-defined phenotype mappings to initialize topic priors, assigning higher weights to phenotype topics with respect to a patient’s observed PheCodes; MixEHR-seed infers expert-guided phenotype representations with a mixture of “seed” topics and regular topics, where a seed is referred to as clinically related ICD codes for a given PheCode. Overall, MixEHR-G employs an expert-guided prior that confines the inferred phenotype topics to pre-defined PheCodes. In contrast, MixEHR-Seed relies solely on expert-guided topic inference, but may face convergence issues due to uninformative initialization. Thus, to leverage the strengths of both approaches, we seek to develop a novel approach that integrates expert knowledge into initialization and adaptively controls expert-guided topic inference through learned weights.

In this study, we present a PheCode-guided, multi-modal topic model called MixEHR-SAGE (Seed-and-Guided). Our approach uses existing disease codings from PheCodes to infer 1,213 phenotype topics, which can leverage complementary information from other EHR data types in a data-driven manner. Specifically, MixEHR-SAGE uses PheCodes to initialize topic priors and infers seed-guided phenotype topic distributions. While MixEHR-SAGE uses PheCodes to guide the topic inference for the ICD codes, unguided modalities such as medication use and medical procedures, which are not captured in ICD codes, still benefit from this information without requiring pre-specified disease definitions. We evaluate MixEHR-SAGE on large-scale EHR datasets, including the UKB and MIMIC-III intensive care unit (ICU) datasets. Our analyses demonstrate that MixEHR-SAGE identifies meaningful disease topics, enhances disease prediction, and discovers novel genetic associations.

## 2 Results

### 2.1 MixEHR-SAGE overview

MixEHR-SAGE consists of 3 key steps: (1) assembling a multi-modal EHR dataset (i.e. ICD codes, medications, procedures, etc.) with modality-specific bag-of-words representations; (2) fitting mixture models to patients’ PheCode counts to estimate topic prior probabilities for each reference phenotype; and (3) inferring the posterior distributions of latent variables using a collapsed variational inference algorithm (Figure 1). In Section 2.2, we first evaluated the quality of phenotype topics inferred by MixEHR-SAGE in the UKB. In Section 2.3, we assessed MixEHR-SAGE phenotype topics’ ability to predict incident diagnoses based on baseline EHR features in the UKB. In Section 2.4, we performed genetic association analyses using derived continuous disease risk scores leading to the idenfitication of novel and meaningful genetic associations.

**Figure 1:**
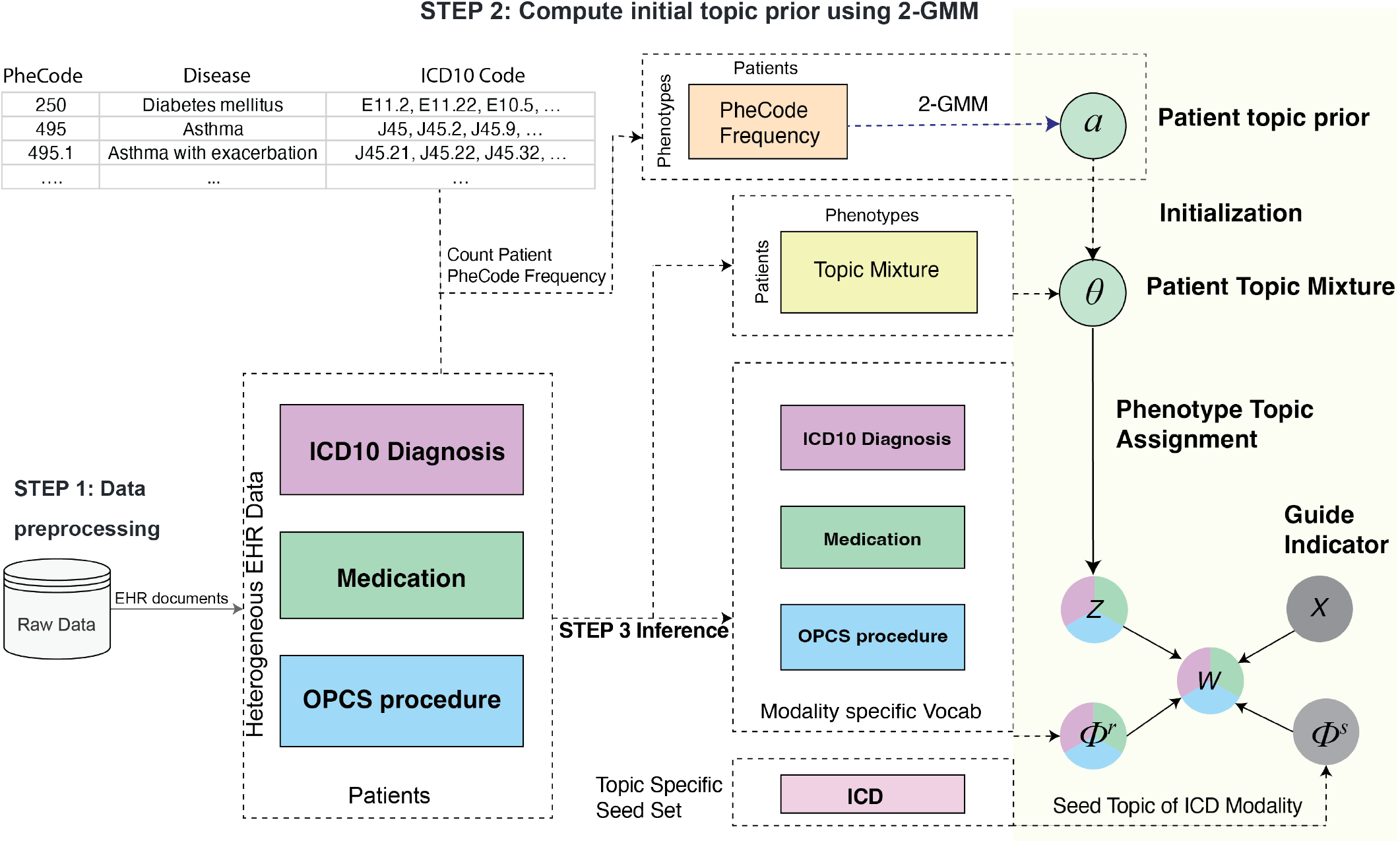
Schematic view of MixEHR-SAGE on the UK Biobank data. In step 1, MixEHR-SAGE ingests patients’ multi-modal EHR data as input. In step 2, MixEHR-SAGE initializes patient topic mixture *θ* by fitting 2-component GMM to PheCode count matrix. In step 3, MixEHR-SAGE performs variational inference to infer latent variables including seed topics 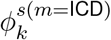 and regular topics 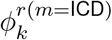 for ICD modality, regular topics for other unguided modalities 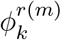, and patient topic mixture *θ. W* represents EHR observations, and *Z* represents the latent topic assignment.

### 2.2 MixEHR-SAGE discovers interpretable topics from UKB data

To ensure the broad applicability of our proposed method, we evaluated MixEHR-SAGE’s ability to capture meaningful phenotype topics from the EHR data in UKB dataset (Figure 1). To validate the interpretability of the inferred disease topics, we extracted the top ICD 10 codes, Anatomical Therapeutic Chemical codes (ATC), and Office of Population Censuses and Surveys (OPCS-4) codes with the highest probabilities for the 6 selected phenotype topics, focusing on two case studies involving diabetic diseases and leukemia cancers, respectively (Figure 2).

**Figure 2:**
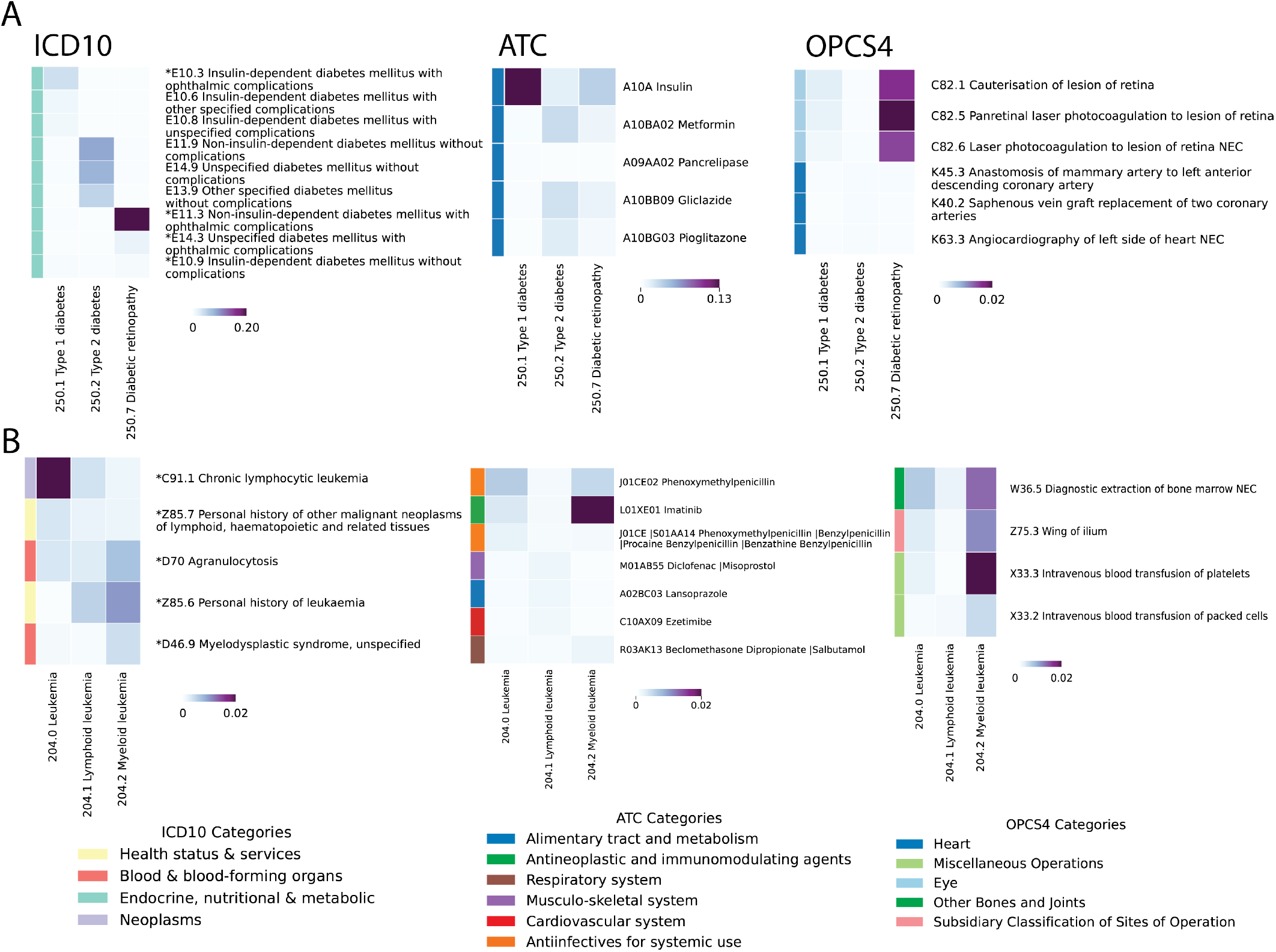
Top 3 highest-probability EHR codes for T2D and leukemia inferred by MixEHR-SAGE. The selected phenotypes belong to diabetes (A) and leukemia (B) categories. For every phenotype, the top 3 EHR codes per modality are presented with duplication removed. For the ICD modality, the ∗ indicate “regular ICD codes” for the corresponding phenotypes. The colorbar represents the probability derived from topic distribution *ϕ*_*dk*_.

The top 3 EHR codes exhibit strong clinical relevance to the selected phenotypes. The topic for Type 1 Diabetes (T1D) includes key insulin-dependent conditions namely “E10.3 insulin-dependent diabetes with ophthalmic complications”, “E10.6 T1D mellitus with other specified complications”, and “E10.8 T1D mellitus with unspecified complications” (Figure 2A). In contrast, the top three ICD-10 codes identified for the Type 2 Diabetes (T2D) topic are “E11.9 Non-insulin-dependent diabetes mellitus”, “E14.9 Unspecified diabetes mellitus”, and “E11.3 Non-insulin-dependent diabetes with ophthalmic complications”, which correspond to different T2D subtypes. For diabetic retinopathy (250.7), “E11.3 Type 2 diabetes mellitus with ophthalmic complications” is the top diagnosis code, which then leads to common microvascular complications, along with E14.3 and E10.9. In the medication modality, the prominent ATC codes are insulin (A10A) for T1D, metformin (A10BA02), the first-line treatment for T2D, as well as gliclazide (A10BB09) and pioglitazone (A10BG03), both of which are commonly used to control blood sugar levels [15, 16]. While diabetes itself is typically managed with medications like insulin or metformin as its first-line therapy, its complications, such as diabetic retinopathy, often require surgical treatments. Indeed, diabetic retinopathy is more strongly linked to ophthalmologic procedures such as laser photocoagulation for retinal lesions (C82.5, C82.6) and cauterisation of leasion of retina (C82.1) than T1D and T2D. These procedures are standard treatments for proliferative diabetic retinopathy, helping to prevent vision loss [17]. T2D was also identified with other complications, such as anastomosis of the artery (K45.3), and saphenous vein graft replacement of two coronary arteries (K40.2). T2D is strongly associated with increased risk of cardiovascular events like coronary artery disease (CAD) [18], which makes angiocardiography crucial for diagnosing and assessing the severity of CAD in diabetic patients [19].

For the leukemia topic 204.0, “C91.1 chronic lymphocytic leukemia”, “Z85.7 history of lymphoid neoplasms”, and “D70 agranulocytosis” are the 3 top ICD 10 codes (Figure 2B). The first two are essentially the diagnostic rule of the cancer. “D70 agranulocytosis” is a condition of severely reduced white blood cells that has been shown to increase infection risk and could potentially develop into leukemia [20, 21]. For the myeloid leukemia topic (204.2), “D46.9 myelodysplastic syndrome” is one of the top ICD codes, which aligns with growing recognition of myelodysplastic syndrome as a pre-leukemic state that progresses to acute myeloid leukemia in roughly 30% of cases [22, 23]. For the ATC medication modality, MixEHR-SAGE highlighted imatinib (L01XE01), the hallmark targeted therapy for chronic myeloid leukemia, alongside diclofenac (M01AB55), a non-steroidal anti-inflammatory drug (NSAID) shown to induce apoptosis in acute myeloid leukemia (AML) cell lines [24, 25]. Besides direct treatment, MixEHR-SAGE also identified phenoxymethylpenicillin (J01CE02), underscoring the routine use of antibiotics to treat infections common in leukemia patients. For the OPCS treatment procedure modality, MixEHR-SAGE identified bone marrow extraction (W36.5), which is important for disease diagnosis. Additionally, intravenous platelet transfusion (X33.2) and packed red blood cell transfusion (X33.2) are used to raise platelet counts and control bleeding [26,27]. Together, these findings demonstrate MixEHR-SAGE’s ability to recover a comprehensive, clinically coherent picture of leukemia care.

To further validate MixEHR-SAGE’s ability to produce interpretable disease topics, we applied it to the MIMIC-III dataset [28]. Besides ICD and medication modalities, MixEHR-SAGE also inferred interpretable phenotype topics across Current Procedural Terminology (CPT), Diagnosis Related Group (DRG), lab tests, and doctor notes. These results demonstrate its ability to leverage diverse EHR modalities for comprehensive phenotype characterization. Further details for MIMIC-III applications are provided in Section 10.3.2, and further information for other disease categories of UKB data are provided in Supplementary Fig S3.

### 2.3 MixEHR-SAGE accurately predicts incident diagnoses from baseline characteristics

To evaluate the predictive performance of the inferred topics for clinically meaningful events, we assessed their ability to predict incident diagnoses using baseline characteristics from the UKB. We analyzed 320,253 individuals using data from their first visit. Prevalent events were the events occurring prior to the recruitment visit, based on hospital inpatient data. In contrast, incident events referred to new inpatient diagnoses recorded after the baseline visit, with no prior record of the matching PheCode before the individual’s baseline date [29]. For controls of each target disease, we applied PheCode exclusion criteria to remove individuals with the related PheCode [8]. We evaluated the top-*K* highest-risk individuals identified by MixEHR-SAGE, with *K* ranging from 10 to 100. For each target disease, we ranked patients based on their predicted risk scores derived from patient topic mixture (i.e., *θ*_*dk*_ in (1)) (Figure 3). We observe that individuals with an incident diagnoses had higher average predicted risk scores than the controls. Using T2D and leukemia as case studies, we identified the top 10 patients (*K* = 10) with the highest predicted risk scores for each disease. We examined their most frequently associated phenotypes, medications, and surgical procedures to assess the key features contributing to model prediction. The predicted risk distributions for incident cases and controls demonstrated a right-skewed pattern among individuals, who later developed T2D and leukemia. Therefore, MixEHR-SAGE can effectively identify high-risk individuals prior to their future clinical diagnosis (Figure 4A, B).

**Figure 3:**
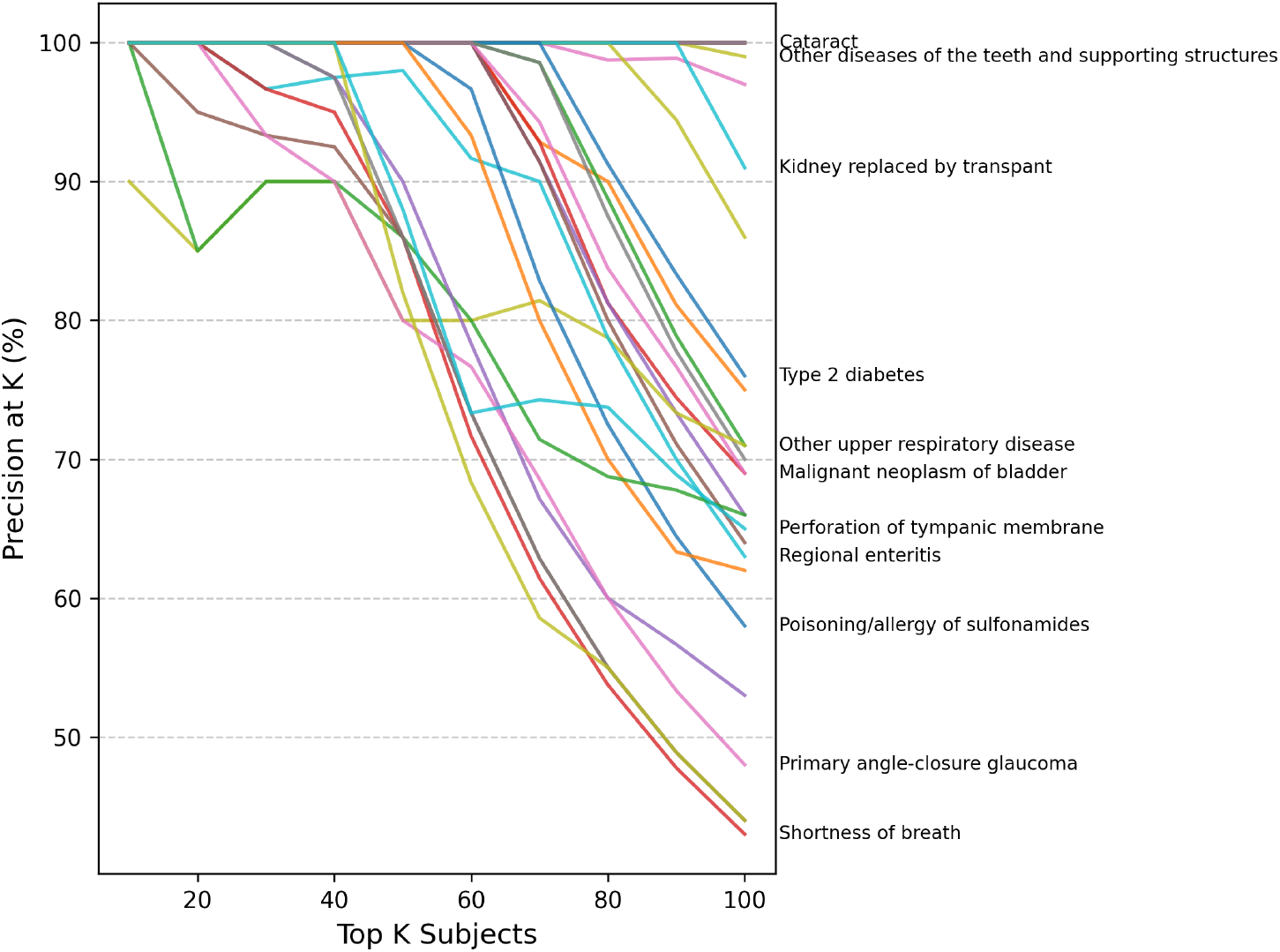
Disease risk prioritization of UKB individuals using patient-level topic mixture inferred by MixEHR-SAGE. Precision as a fraction of number of incident cases among top *K* patients, where *K* ∈ {10, …, 100}. Each line represents the change in precision for a disease as K increases. Notably, some diseases, such as T2D and cataracts, maintain high precision across *K*-values.

**Figure 4:**
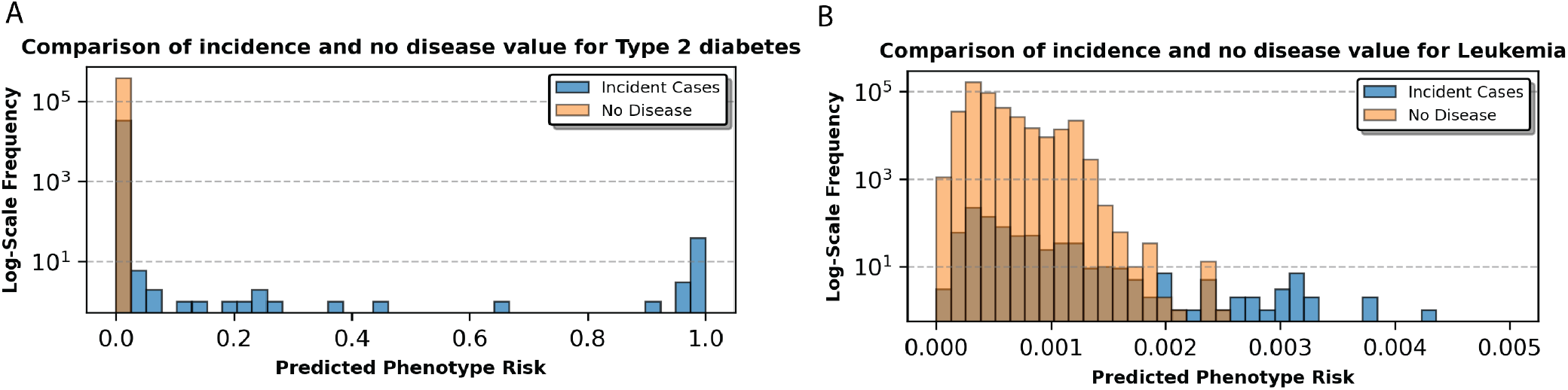
Predicted phenotype risk and clinical features for T2D and leukemia case studies. The Bar plots compare the prevalence (y-axis) of the predicted phenotype risk scores (x-axis) between incident cases (blue) and the control group (orange) for (A) T2D and (B) leukemia.

To evaluate the clinical relevance of the estimated T2D risk scores, we analyzed high-risk individuals identified by MixEHR-SAGE before and after the baseline assessment. Concordant to the above topic analysis (Section 2.2), among those high-risk individuals, some patients had already been diagnosed with T2D-related complications (e.g., diabetic retinopathy), prescribed antidiabetic medications (e.g., insulin, metformin), and received relevant procedures associated with diabetic retinopathy like C82.1 and C86.5. (Figure 5A). Although these individuals were not T2D cases based on PheCodes before the baseline visit, our model assigned them high-risk scores for the T2D topic, demonstrating its utility for early diagnosis. After the baseline visit, individuals were diagnosed with T2D, either with or without complications (E11.3, E11.9). Notably, a significant number of patients were also diagnosed with essential hypertension (I10), a common condition in the UKB population and a well-known comorbidity of T2D (Figure 5A right). Newly diagnosed procedures, such as phacoemulsification of the lens (C71.2), were associated with diabetic retinopathy and also with patients diagnosed with T2D and ophthalmic complications. A recent study reviewed the safety of this surgery for T2D patients, and it is considered a safe treatment with careful management [30].

**Figure 5:**
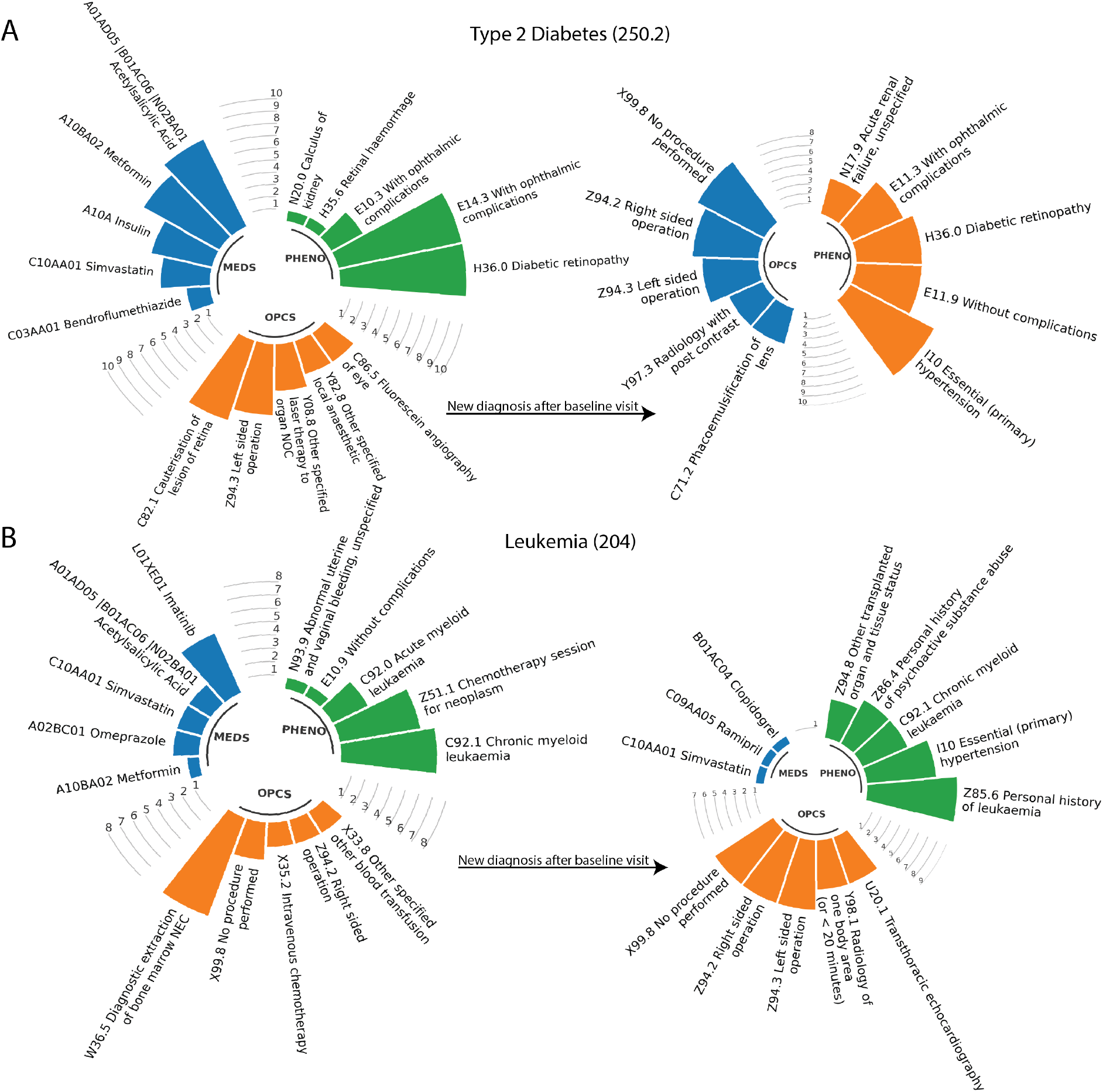
Top k patients and their most frequent EHR codes before and after baseline visits. The circular barplot illustrates the top 5 most frequent EHR codes from the Medications (MDES), PheCodes (PHENO), and OPCS modalities observed among the top 10 high-risk patients inferred by MixEHR-SAGE before and after their baseline visits. (A) T2D and (B) Leukemia.

For Lymphoid Leukemia (PheCodes 204), many high-risk individuals identified by MixEHR-SAGE already exhibited pre-existing conditions related to leukemia before the baseline visit. Risk estimation was primarily driven by the occurrence of leukemia-related phenotypes and procedures (Figure 5B left panel). In particular, procedures such as diagnostic extractions of bone marrow, also refers to as bone marrow biopsies (W36.5) and intravenous chemotherapy (X35.2) suggest that these individuals had prior hematologic abnormalities or were evaluated for related conditions through surgical procedures [31]. Some of the high-risk patients of Lymphoid Leukemia were diagnosed with Chronic Myeloid Leukemia (ICD code C92.1) either at the baseline visit or at the follow-up visit, which maps to PheCode 204.22 rather than 204 (Figure 5B right panel). These results demonstrate MixEHR-SAGE’s ability to capture both broad hematological disease categories and specific leukemia subtypes. Additionally, some individuals in the high-risk group were later diagnosed with a different leukemia subtype, highlighting MixEHR-SAGE’s ability to capture shared risk factors across various hematologic malignancies.

In summary, T2D prediction was strongly driven by medication use and pre-baseline diagnostic codes, resulting in a distinct separation between incident cases and non-disease individuals (Figure 5B). In contrast, leukemia prediction was primarily influenced by pre-existing hematological conditions, resulting in more overlap in risk distributions between incident cases and non-disease individuals (Figure 5A). Overall, MixEHR-SAGE accurately identified high-risk individuals and enabled risk stratification by capturing prevalence and incidence clinical events from the multi-modal EHR data.

### 2.4 MixEHR-SAGE identifies novel genetic associations

Building on the incidence prediction results, we conducted GWAS across 525 phenotypes, each of which has greater than 1000 cases, using both the traditional binary disease labels and our continuous disease risk scores inferred from MixEHR-SAGE. The majority of genome-wide significant loci were identified by both approaches, and MixEHR-SAGE further uncovers novel loci in several phenotypes (Figure 6).

**Figure 6:**
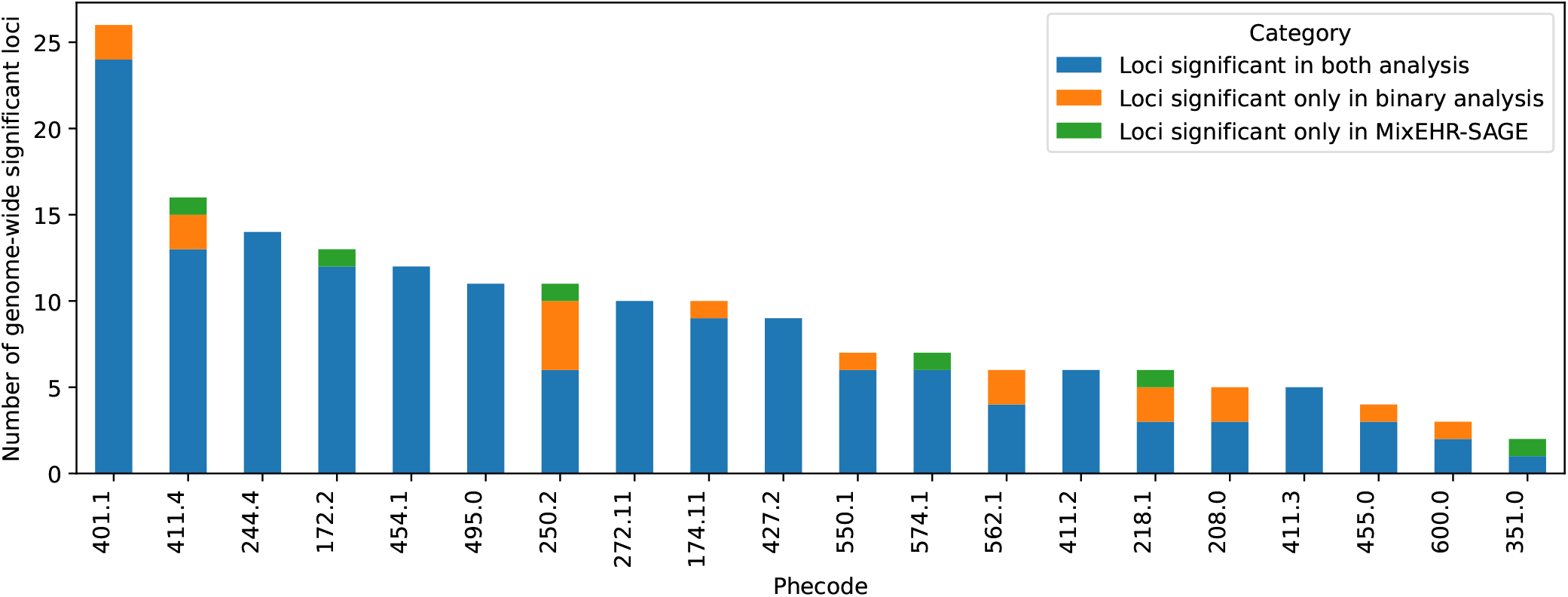
Summary of the number of loci reaching genome-wide significance (*p <* 5×10^−8^) in the binary Phecode and MixEHR-SAGE models. The 20 most common diseases according to the Phecode definitions (i.e. most prevalent) are shown, sorted by the total number of identified loci. The loci are defined using LD-based clumping. The 6 phenotypes (and their Phecodes) with the largest number of genome-wide significant loci detected using MixEHR-SAGE are: Coronary Atherosclerosis (411.4; n loci = 16), Other non-epithelial cancer of skin (172.2; n loci = 13), Type 2 Diabetes (250.2, n = 7), Cholelithiasis (574.1; n loci = 7), Uterine leiomyoma (218.1; n loci = 4), and Other peripheral nerve disorders (351; n loci = 2).

To showcase MixEHR-SAGE’s utility across phenotypes with different prevalence counts, we chose two contrasting examples: T2D and Leukemia. T2D ranks among the top 20 most common diagnoses in UKB. In contrast, leukemia (204) has only 178 cases in UKB but emerged as one of the top 10 phenotypes that show the greatest gain in its expected sample size (*n* = 221). The comparison of genome-wide significant loci identified by MixEHR-SAGE and the traditional binary GWAS for the 100 most prevalent diseases is provided in Supplementary Table S3.

For T2D, we identified a locus harboring *PPP1R15A*, which was missed by the binary approach (Figure 7A, B, C). The lead variants of the locus is rs610308, a previously identified T2D variant [3]. The rs610308 variant is a missense variant of *PPP1R15A* (Protein Phosphatase 1 Regulatory Subunit 15A) based on dbSNP [32]. This gene plays a critical role in cellular stress responses and insulin signalling pathways, and is essential for metabolic regulation and pancreatic beta-cell function [33, 34]. This demonstrates that MixEHR-SAGE can identify genetic variants associated with both T2D and its complications (e.g., diabetic kidney disease). In addition, we also identified two more well-established T2D loci that were missed by the binary model, including *HNF1B* and *CDKAL1* [35, 36] (Figure 7B).

**Figure 7:**
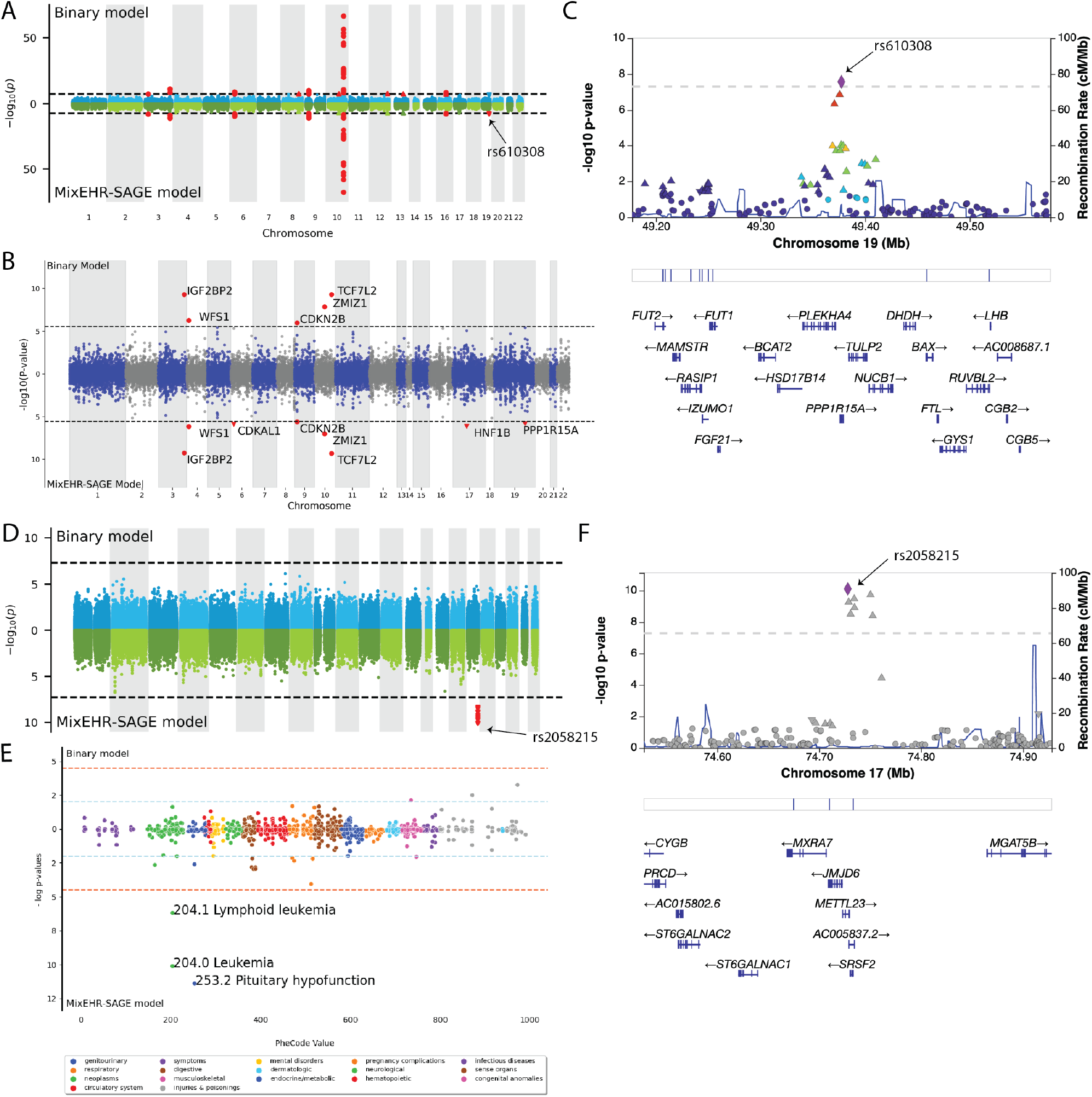
Genetic signals linked to T2D (A-C) and leukemia (D-F) using MixEHR-SAGE topics and PheCode labels. (A) Genome-wide comparison of genetic associations for T2D based on binary PheCodes (top) and MixEHR-SAGE topics (bottom), revealing new findings on chromosome 19. (B) Gene-level analysis of T2D GWAS showing the significantly associated genes (*P* ≤ 0.05*/*17549 = 2.849^−6^). (C) A zoomed-in plot of chromosome 19 of a T2D locus showing the location of the significant loci and its nearby genes. (D) Genome-wide comparison of genetic associations for leukemia using binary PheCode labels (top) and MixEHR-SAGE-inferred topics (bottom). MixEHR-SAGE successfully identified new loci on the chromosome 17. (E) The phenome-wide association analysis for the leukemia locus harboring genetic variant rs2058215. This reveals significant association of 2 types of leukemia, namely lymphoid leukemia and leukemia. (F) Locus Zoom plot of the rs2058215-locus, showing the significant hits and its closest genes.

To replicate our findings independently, the T2D-associated loci identified by our approach are compared to those reported in the multi-ancestry meta-analyses of Suzuki et al. [37]. All identified loci were successfully replicated in this large study when using windows defined from LD clumped lead SNPs using the method described in Section 3.6. In some instances, the lead variants appeared to differ between the two approaches (Supplementary Fig. S4). This is likely due to differences in LD patterns between the ancestral backgrounds present in our cohort and the multi-ancestry meta-analysis panel, or due to differences in the statistical power to detect specific variants.

Importantly, the fact that our MixEHR-SAGE infers continuous phenotype probabilities for each individual enables performing GWAS on some of the low-prevalent phenotypes such as leukemia, for which the number of cases are fewer than 1000. As a result, the traditional binary phecode-based GWAS analysis is unable to detect any GWAS locus due to the lack of power. On the other hand, our approach identified a significant locus, which was missed by the conventional binary method (Figure 7D). This association signal was led by the leading SNP rs2058215. When performing PheWAS on this leading SNP, MixEHR-SAGE also indicated its association with lymphoid leukemia (204.1) (Figure 7E). The locus harbors *JMJD6, METTL23*, and *MFSD11*, as well as a 3’ UTR variant of *SRSF2* (Figure 7F). The gene *JMJD6* is implicated in various cancers, including leukemia, due to its versatile role in epigenetic regulation, RNA splicing, and histone modification [38]. It contributes to tumor development mainly through regulating immune responses and cancer-related pathways. It also plays a crucial role in the differentiation of hematopoietic stem cells, a critical process in leukemia pathogenesis [38]. *SRSF2* (Serine and arginine-rich splicing factor 2) is known to regulate gene expression essential for normal hematopoiesis. Mutations in *SRSF2* have been strongly associated with various myeloid malignancies, including acute myeloid leukemia (AML), myelodysplastic syndromes (MDS), and chronic myelomonocytic leukemia (CMML) [39, 40]. Functional studies show that SRSF2 mutations alter hematopoietic stem cell differentiation, leading to clonal expansion of preleukemic cells. Furthermore, *SRSF2* mutations frequently co-occur with mutations in epigenetic regulators such as *TET2, ASXL1*, and *DNMT3A*, suggesting a synergistic effect in leukemic transformation [41–43]. Given its key role in RNA processing and leukemia pathogenesis, *SRSF2* is being increasingly explored as a therapeutic target, with current efforts focusing on small molecules that modulate RNA splicing [44]. In addition, rs2058215 is in high LD (*r*^2^ = 0.69) with rs9897202, an eQTL SNP for *MXRA7* based on whole blood samples from GTEx [45]. *MXRA7* is involved in hematopoiesis and immune response and serves as a possible effector gene on leukemia risk [46]. A recent study demonstrated that *MXRA7* could influence the pathogenesis of acute promyelocytic leukemia (APL) through regulation of cell differentiation [47].

Together, by modeling disease risk on a continuum via MixEHR-SAGE, we capture genetic signals often missed by the conventional GWAS that relies on the binary disease labels.

## 3 Materials and Methods

### 3.1 MixEHR-SAGE details

We consider each patient’s medical records as a document indexed by *d* ∈ {1, …, *D*}. Each patient’s phenotype topic mixture *θ*_*d*_ is generated from a *K*-dimensional Dirichlet distribution with a prior parameter *α*. For a document of size *N*_*d*_, each EHR observation (*e.g*., a ICD code) is treated as a word 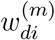, where *i* ∈ {1, …, *N*_*d*_} indexes the word and *m* ∈ {1, …, *M*} indicates the data modality. Each word 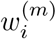 is associated with a latent topic assignment *z*_*di*_ ∈ {1, …, *K*}.

MixEHR-SAGE incorporates expert-defined PheWAS mapping as domain knowledge to guide topic inference and improve clinical interpretability. Specifically, PheWAS projects each PheCode to a set of clinically related ICD codes, allowing us to infuse phenotype concepts directly into MixEHR-SAGE. As a result, MixEHR-SAGE can learn over 1,000 phenotype topics, each corresponding to an expert-defined phenotype concept. For the expert-guided ICD modality, MixEHR-SAGE utilizes seed-guided topic mechanism [14] to represent each phenotype topic *k* ∈ {1, …, *K*} with two distributions: a seed-topic distribution 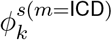 over only its seed set 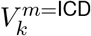 and a regular-topic distribution 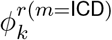 over the entire feature vocabulary *V* ^*m*=ICD^. The seed topics capture the phenotype-the specific expert knowledge, while the regular topics govern the general phenotype distributions. Each ICD code 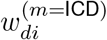 is drawn from a mixture distribution: 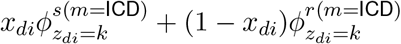, where *x*_*di*_ indicates whether an ICD code 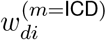 is generated from a seed topic (*x*_*di*_ = 1) or a regular topic (*x*_*di*_ = 0). The indicator *x*_*di*_ follows a Bernoulli distribution with the seed-topic rate *π*_*k*_, representing the probability that an ICD code is drawn from the *k*-th seed topic.

In contrast, a non-ICD EHR code 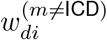 (e.g., medications, procedures) is only sampled from the regular topic 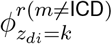. Although these unguided modalities do not directly benefit from the incorporated expert knowledge, MixEHR-SAGE shares PheCode-guided information through the patient topic mixture *θ*.

The implementation of the proposed MixEHR-SAGE model comprises three key steps:

1. **Multi-modal EHR Data Construction**: We processed the multi-modal EHR dataset comprising ICD codes, medications, and OPCS-4 procedures into 3 matrices (Figure 1 step 1). A detailed description of the UK Biobank data processing pipeline is provided in Section 3.2.

#### Algorithm 1 Inference algorithm of MixEHR-SAGE

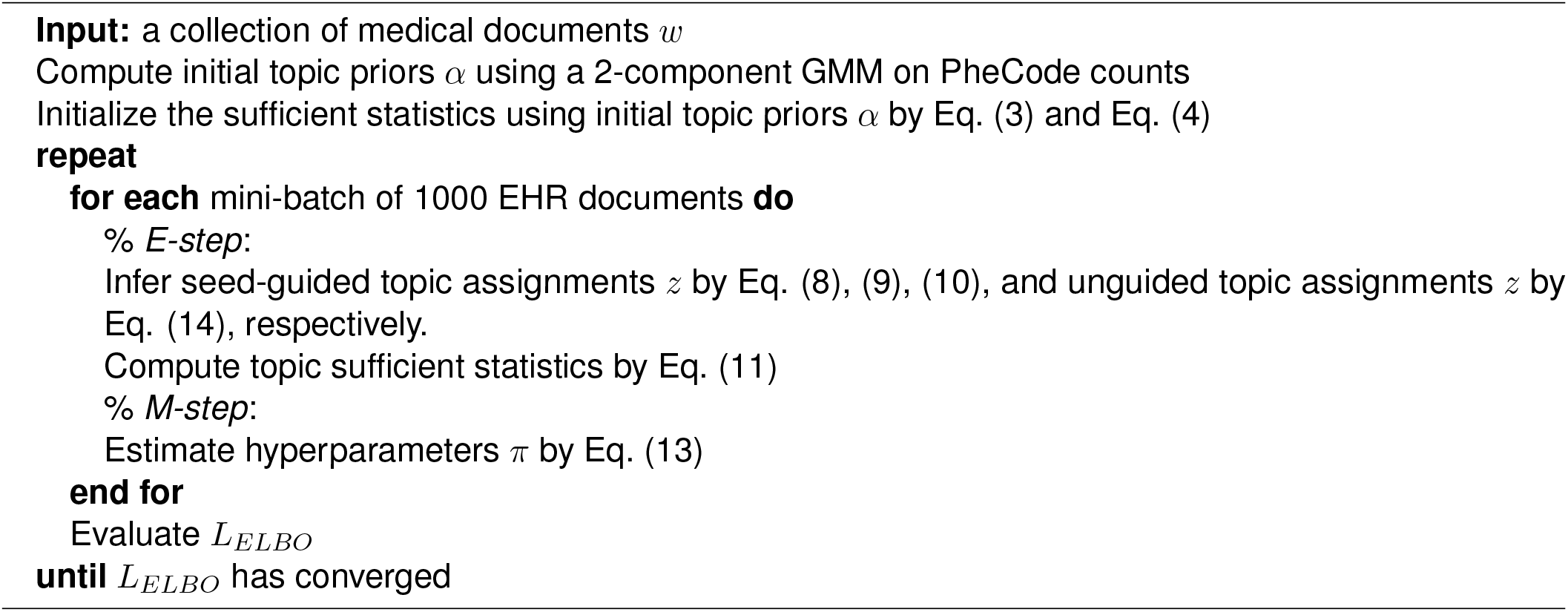
2. **Initialization of Pheontype Topic Priors**: For each phenotype, we fit a two-component Gaussian mixture model (GMM) to patients’ PheCode counts to estimate the topic prior *α*_*k*_ (Figure 1 step 2). Further details on this procedure are provided in Section 10.2.1.
3. **Posterior Approximate Inference**: We infer the posterior distributions of latent variables using a mean-field, collapsed variational inference algorithm (Figure 1 step 3). The full mathematical formulation and derivation of this inference algorithm are provided in Section 10.2.2.

The complete inference algorithm is summarized in Algorithm 1. To effectively handle large-scale EHR data, we performed stochastic variational inference using mini-batches containing 1,000 EHR documents per batch [48]. We used the validation set to fine-tune the topic hyperparameters *µ* and *β* to minimize the held-out negative log-likelihood. Upon convergence of the ELBO, we can compute the collapsed variables (*θ, ϕ*^*r*^, *ϕ*^*s*^) with their respective variational expectations:

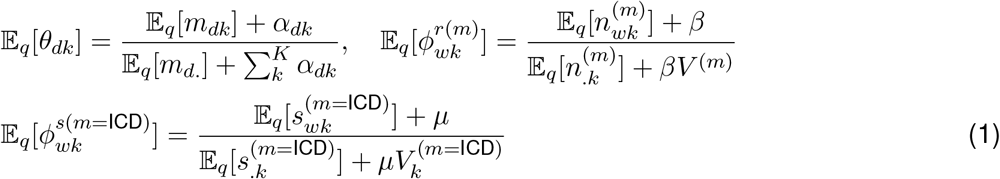

Here, *m*_*dk*_ denotes the sufficient statistic for the topic mixture of patient *d*, and *m*_*d*·_ is its summation over all topics. For sufficient statistic of topic assignments, *n*_*wk*_ indicates the number of times word *w* is assigned to regular topic *k*, and *s*_*wk*_ indicates the number of times of all seed word *w* is assigned to seed topic *k*. Additionally, *n*_·*k*_ and *s*_·*k*_ are the total counts of regular words and seed words assigned to topic *k* across all patients, respectively. The notations of variables are defined in Table S2. The full derivation of the inference algorithm is provided in Section 10.2.2.

### 3.2 UK Biobank data processing

The UK Biobank is a prospective population cohort comprising approximately 500,000 participants aged 40 to 69 at recruitment [49]. At the recruitment visit, participants provided informed consent and answered questions on socio-demographic and health-related factors, including medication use. Hospitalization data was obtained through the Health Episode Statistic (HES) dataset, with records included up to January 2022. To enable large-scale genomic analysis, genome-wide genetic variants were assessed in all participants using the UKB Axiom Array, which measures approximately 850,000 variants. Genotype imputation procedure using the Haplotype Reference Consortium, the UK10k and the 1000 Genomes Consortium as a haplotype reference panel allowed over 90 million variants to be imputed.

An overview of EHR data preprocessing and quality control is provided in Figure 8. We utilized three data modalities from the UKB to train MixEHR-SAGE. We included ICD-10 diagnostic codes corresponding to both primary and secondary causes of hospitalization. We also included self-reported medication codes (variable #20003), collected during verbal interviews with a research nurse conducted during any assessment center visit to capture pharmacotherapy data as a second modality. As the third modality, we incorporated OPCS-4 codes capturing surgical and operative procedures. We excluded any EHR code with less than 10 occurrences. We mapped 6,745 unique drug codes in UKB to 885 ATC codes using a published mapping method, grouping related drugs together into clinically meaningful therapeutic categories [50]. The processed UKB data included 6,954 ICD-10 codes and 2,560 OPCS-4 codes.

**Figure 8:**
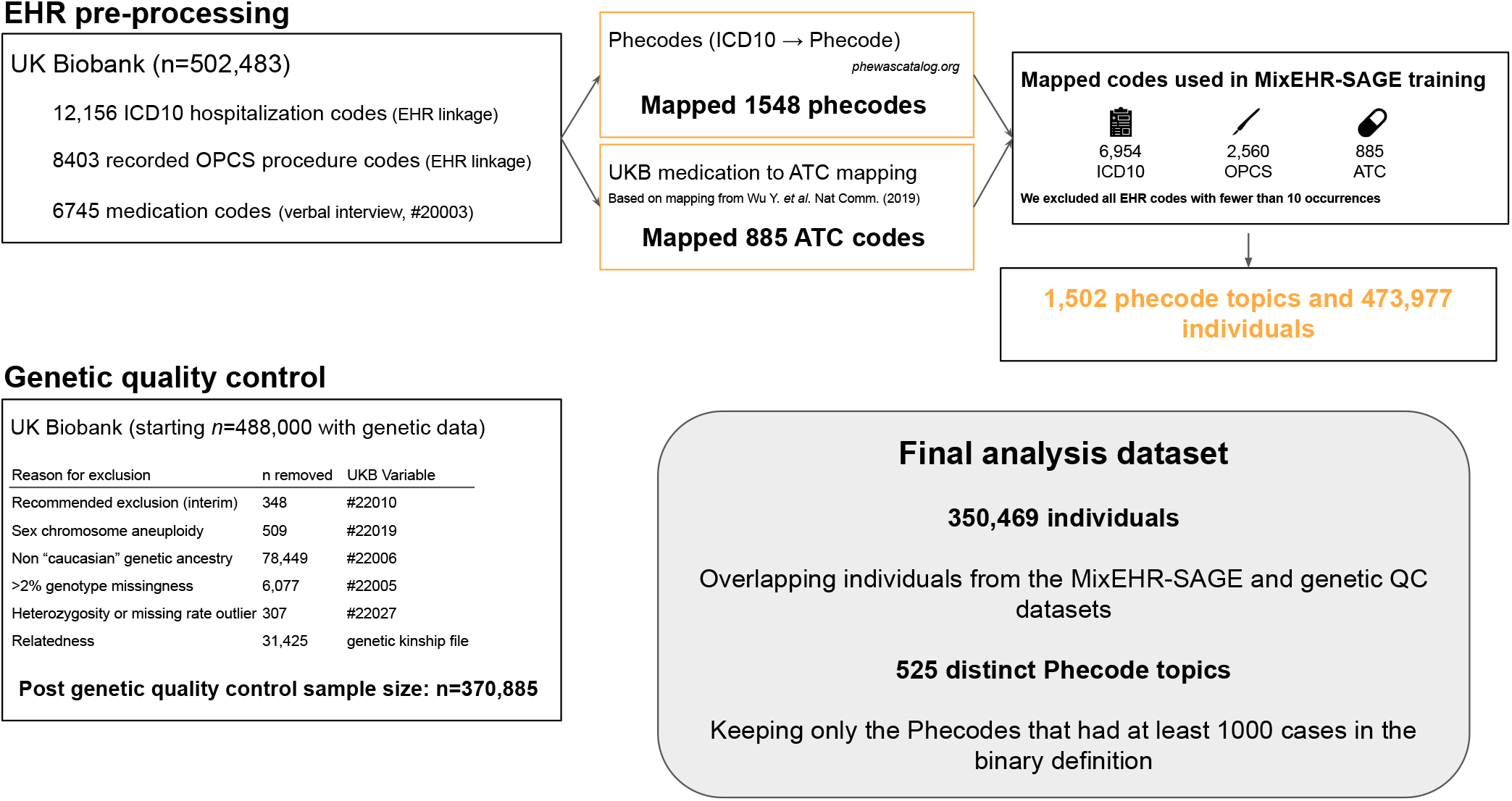
Overview of the UKB data quality control steps used to train MixEHR-SAGE. Following exclusion criteria during genetic quality control, including non-European ancestry, excess missing rate, and relatedness.

### 3.3 Inferring expected phenotypes

The probability of disease incidence was calculated based on the generative process of latent Dirichlet allocation (LDA) [51]. Specifically, we inferred the expected patient-by-ICD matrix by multiplying the patient topic mixture *θ* ∈ ℝ^*D×K*^ with the topic distribution matrix 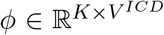. The inferred phenotype probabilities are at low numerical scale because it is derived from Dirichlet distribution with dimensionality equal to the total number of phenotypes (i.e., over 500). To be comparable with the binary phenotype and also improve GWAS analysis, we sought to rescale the inferred phenotype probabilities while maintaining the relative difference among individuals. Specifically, for each inferred ICD code, we fit a simple logistic regression treating the binary ICD code *j* for patient *d* as the response variable and the inferred ICD probabilities as the input feature:

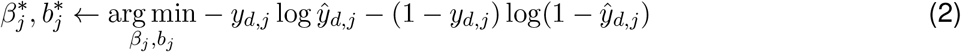

where 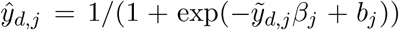 and 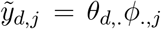. We then aggregated ICD codes onto their corresponding PheCodes, resulting in a Patient-by-PheCode matrix with 1,213 unique PheCodes. The expected sample sizes for each PheCode *k* ∈ {1, …, *K*} under the new matrix are calculated by summing its columns across all patients *D*.

### 3.4 Genetic quality control

Prior to performing GWAS, we conducted genotype quality control (QC) to ensure the reliability of the genetic data. This involved applying several filtering steps to exclude individuals based on genotyping quality metrics, ancestry, and relatedness (Figure 8).

We first excluded individuals marked as Recommended Genomic Analysis Exclusions (variable #22010, *n* = 348) due to poor heterozygosity or high missing rate. Additionally, we excluded those with sex chromosome aneuploidy (variable #22019, *n* = 509). To account for population stratification, we excluded individuals of non-European genetic ancestry identified by genotype-based Principal Component (PC) Analysis (variable #22006, *n* = 78,449). We further excluded individuals with more than 2% genotype missingness (variable #22005, *n* = 6,077), as high levels of missing data may indicate poor genotyping quality. We additionally removed individuals identified as outliers in heterozygosity and missingness rate (variable #22027, *n* = 307), as substantial deviations from the expected distribution may reflect genotyping errors or sample contamination. Finally, because logistic regression assumes independent samples, we excluded related individuals to prevent statistical inflation of from genetic relatedness. Specifically, we removed one individual at random from each pair with a kinship coefficient greater than 0.0884 (variable #22021, *n* = 31,425). After applying all genetic QC criteria, the final dataset included 370,885 unrelated individuals of European ancestry with high-quality genotyping data, ensuring a robust foundation for association analysis.

### 3.5 Association analysis

To assess genetic associations across phenotypes, we performed GWAS using PLINK 1.9 [52]. We applied two statistical models for GWAS: (1) logistic regression using binary phenotypes based on PheCode definitions, adjusting for age, sex, and 20 principal components (PCs) as covariates; and (2) linear regression using continuous disease risk scores inferred from MixEHR-SAGE while adjusting for the same covariates. For both models, we applied standard GWAS QC procedures described in Section 3.4. To define independently associated genetic loci, we used linkage disequilibrium (LD) clumping as implemented in PLINK v1.9. We clumped variants based on a primary significance threshold of *p <* 5 *×* 10^−8^, a clumping window of 1000kb around each index SNP, and an LD cutoff of *r*^2^ = 0.2.

### 3.6 External Validation

To perform external validation, we compared our T2D-associated loci to those reported by Suzuki [37], a large-scale T2D meta-analysis including 428,452 cases from the Type 2 Diabetes Global Genetics Initative (T2DGGI). Lead SNPs for independent loci were defined separately in our UKB analysis and the European subset of the T2DGGI study via the same LD-based clumping method 3.5. Padding regions of *±*500kb were added to the lead SNPs to compare significantly-associated loci from both studies using bedtools 2.31.0 [53]. For visualization of regional association signals, we generated locus-specific plots using LocusCompare, which facilitates side-by-side comparison between the two studies while incorporating LD structure from the 1000 Genomes EUR reference panel [54].

## 4 Discussion

In this study, we introduce MixEHR-SAGE, a seed-guided topic model designed to improve the phenotypic representation and discovery of novel GWAS loci. By integrating PheCode-guided prior knowledge, MixEHR-SAGE enables the inference of more than 1,000 interpretable phenotype topics, each aligned with clinically defined disease concepts. A key property of MixEHR-SAGE is its ability to integrate multiple data modalities (e.g., ICD codes, medications, and procedures), thereby capturing more comprehensive and clinically meaningful disease representations. Unlike traditional phenotyping methods that solely rely on binary case-control definitions, MixEHR-SAGE models diseases as continuous risk scores, reflecting the spectrum of phenotype severity. This approach is particularly beneficial for diseases with varying severity levels, subtypes, or progression stages, such as metabolic disorders and cancer.

We applied MixEHR-SAGE to the UKB dataset, demonstrating its ability to infer interpretable phenotype topics and improve GWAS discovery. The inferred topics are highly interpretable even for modalities without explicit expert knowledge, such as medications and procedures. We then utilized each patient’s topic mixture as continuous disease risk scores to predict incident disease onset. MixEHR-SAGE accurately identified high-risk individuals for developing T2D and leukemia. For T2D, MixEHR-SAGE captured early pharmacotherapy signals such as metformin prescriptions; for leukemia, relevant ICD diagnostic and OPCS procedure codes were leveraged. Through GWAS analysis, we identified novel genetic signals complementary to those found by traditional binary models. For T2D, MixEHR-SAGE revealed genetic associations specifically linked to clinically relevant complications such as diabetic retinopathy and diabetic kidney diseases. For leukemia, MixEHR-SAGE identified a novel loci harboring *JMJD6* that was missed by the binary approach.

There are several future directions to be explored. MixEHR-SAGE primarily relies on categorical EHR data. However, many additional clinical data types, such as laboratory tests, biosignals (electrocardiography), and images(e.g. MRI), are inherently continuous. In addition, the categorical EHR data can also be influenced by non-biological factors such as age, socioeconomic status (SES), healthcare utilization, and follow-up time. As the UKB continues to accrue medical information such as longitudinal medical records and lifestyle factors, MixEHR-SAGE can be further extended to capture evolving disease trajectories. Future research can extend MixEHR-SAGE to incorporate continuous-valued EHR data, further improving phenotype modelling and genetic discovery.

Another potential limitation relates to the uncertainty of the continuous risk scores inferred by MixEHR-SAGE. While the model produces posterior distributions over patient-topic proportions we used point estimates for downstream risk scores and GWAS analyses in the current study. Recent work demonstrates that incorporating uncertainty-aware phenotypes into downstream analyses can improve robustness and interpretability, and similar approaches could be integrated into MixEHR-SAGE as future work [55].

Unlike purely unsupervised models, which can identify *de novo* phenotype clusters without predefined labels, MixEHR-SAGE uses PheCode-seeded priors to guide topic inference. This improves interpretability and ensures alignment with known disease categories. Although MixEHR-SAGE topics can expand or deviate from their initial seeds, wholly novel clusters are less likely to emerge compared to unsupervised models. Future research could explore a hybrid approach, allocating a portion of topics for unguided discovery while seeding others for interpretability, similar to MixEHR-Nest, which is a hierarchical guided-topic modeling approach to infer subphenotypes from multi-modal EHR data. [56].

A key challenge in genetic studies is its restricted generalizability across diverse populations. Although the UKB dataset includes a diverse cohort, it predominantly consists of individuals of European ancestry, limiting the generalizability of our findings to multi-ethnic populations. Since genetic architecture and disease prevalence vary by ancestry, future work could extend MixEHR-SAGE to more diverse cohorts. Additionally, we excluded rare diseases due to limited sample sizes, and future work could explore adapting MixEHR-SAGE for low-prevalence conditions. Although MixEHR-SAGE demonstrated improved genetic associations, it did not explore polygenic risk score (PRS) prediction. Future work could explore novel PRS models using probabilistic phenotype representations that reflect continuous disease severity rather than binary case definitions.

Another direction for future research is incorporating longitudinal modeling and survival analysis into MixEHR-SAGE. While MixEHR-SAGE effectively stratifies disease phenotypes, it does not explicitly model disease progression and predict survival outcomes. Many chronic diseases, including cancer, cardiovascular conditions, and neurodegenerative disorders, exhibit dynamic progression patterns that could be better captured by time-to-event models. Incorporating survival analysis methods (e.g., Cox proportional hazards models or deep learning-based survival models) into MixEHR-SAGE framework could enhance phenotype risk stratification for chronic diseases. Recent advancements like MixEHR-SurG, which demonstrated promising results on Quebec CHD and MIMIC-III datasets, could be used to extend MixEHR-SAGE framework to analyze survival outcomes in the UKB dataset [57]. Additionally, by integrating the inferred phenotype topics with Mendelian Randomization (MR) could help identify causal relationships between genetic variants and disease progression mechanisms. Lastly, federated learning or privacy-preserving EHR modeling is another emerging challenge/opportunity [58].

In summary, our results demonstrate the value of probabilistic topic modeling for multi-modal EHR phenotyping to improve GWAS discovery. MixEHR-SAGE offers a scalable framework capable of phenotyping over 1,000 phenotypes and identifying novel genetic associations beyond traditional binary case definitions. As biobanks expand, more diverse data types such as biosignals, exome sequencing, and whole-genome sequencing will become available. Automatic phenotyping approaches such as MixEHR-SAGE are critical for integrating clinical phenotyping with genetic discovery, thereby advancing biomedical informatics and genetic epidemiology.

## 5 Data availability

The UKB data used in this study are available to bona fide researchers upon application http://www.ukbiobank.ac.uk/. Derived data supporting the findings of this study are available from the corresponding author upon reasonable request.

## 6 Code availability

The custom code and scripts used to implement MixEHR-SAGE, including sample data preprocessing and topic modeling pipelines, are publicly available at GitHub via the following link: https://github.com/li-lab-mcgill/MixEHR-SAGE.

## 7 Acknowledgments

Y.L. was supported by the following funding programs: the Canada Research Chair (Tier 2) in Machine Learning for Genomics and Healthcare (CRC-2021-00547), Natural Sciences and Engineering Research Council (NSERC) Discovery Grant (RGPIN-2016-05174) and NOVA-FRQNT-NSERC grant (FRQ-NT 2023-NOVA-328677). This research used the NeuroHub infrastructure and was undertaken thanks in part to funding from the Canada First Research Excellence Fund, awarded through the Healthy Brains, Healthy Lives initiative at McGill University. This research was enabled in part by support provided by Calcul Québec and the Digital Research Alliance of Canada. This research has been conducted using the UK Biobank Resource under Application Number 45551.

## 8 Author contributions

Y.L. conceived the study. Y.L. and M.A. supervised the work. Z.S. implemented the method. Z.Y. processed the UKB data and conducted the analysis with help from S.Z. and M.A.. All authors edited and reviewed the final version.

## 9 Declaration of Interests

The authors declare no competing interests.

## Supplementary Information

### 10 Supplementary Methods

#### 10.1 Methodological background

##### MixEHR-Seed model

MixEHR-Seed performs seed-guided topic modeling by computing each phenotype topic *k* of ICD modality with two distributions: a seed-topic distribution 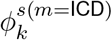 over only its seed set 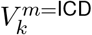 and a regular-topic distribution 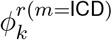 over the entire feature vocabulary *V* ^*m*=ICD^. The regular topics govern the global phenotype distributions and the seed topics capture the phenotype-specific information.

Each ICD code 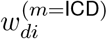 is drawn from a mixture distribution: 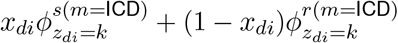, where *x*_*di*_ indicates whether an ICD code 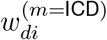 is generated from a seed topic (*x*_*di*_ = 1) or a regular topic (*x*_*di*_ = 0). The seed-topic rate *π*_*k*_ controls the sampling probability that an ICD code is drawn from the seed topic rather than the regular topic. In contrast, for other unguided modalities, a non-ICD EHR feature 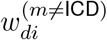 is only sampled from the regular topic 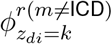. Although other unguided modalities do not directly benefit from expert knowledge, MixEHR-Seed shares guided information through the general patient topic mixture variable *θ*_*d*_.

In summary, the MixEHR-Seed EHR data generative process is described as follows:

1. For each phenotype topic *k* = {1, …, *K*} of ICD modality *m* = ICD:
  a. Draw regular topic 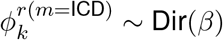 over the entire ICD vocabulary *V* ^*m*=ICD^
  b. Draw seed topic 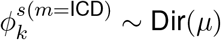 over only the topic-specific seed set 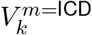
  c. Draw seed-topic rate *π*_*k*_ ∼ Beta(1, 1)
2. For each phenotype topic *k* = {1, …, *K*} of other unguided modalities *m*≠ ICD:
  a. Draw regular topic 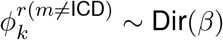 over the entire feature vocabulary *V* ^(*m*≠ICD)^
3. For each EHR document *d* = {1, …, *D*}:
  a. Draw phenotype topic proportion from topic prior *θ*_*d*_ ∼ Dir(*α*)
  b. For each EHR observation *i* = {1, …, *N*_*d*_}, if 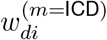 is an ICD code:
    i. Draw topic assignment *z*_*di*_ ∼ Mult(*θ*_*d*_)
    ii. Draw seed-topic indicator 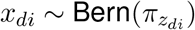.
    iii. Draw an ICD code:

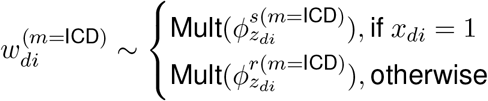
  c. For each EHR observation *i* = 1, …, *N*_*d*_ under other unguided modalities *m*≠ ICD:
    i. Draw topic assignment *z*_*di*_ ∼ Mult(*θ*_*d*_)
    ii. Draw an EHR observation:

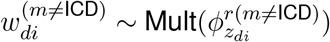

where Dir, Multi, Bern abbreviate Dirichlet, Multinomial, and Bernoulli distributions, respectively.

##### MixEHR-Guided model

MixEHR-G assumes patient topic mixture *θ*_*d*_ is generated from Dirichlet distribution with *K*-dimensional asymmetric hyperparameters *α*_*d*_ = (*α*_*d*1_, …, *α*_*dK*_), which is initialized by a modified Multimodal Automated Phenotyping (MAP) approach [59]. For each phenotype topic *k*, it estimates prior probabilities by fitting Poisson and Lognormal mixture models to *D*-dimensional patients’ PheCode counts. Consequently, a higher prior likelihood of phenotype presence should generate a proportionally higher probability of feature attribution to that phenotype.

In contrast to MixEHR-Seed, MixEHR-G generates phenotype topics solely based on the regular topic distributions and draws EHR feature 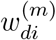 from these regular topics 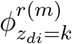.

1. For each phenotype topic *k* = {1, …, *K*} of any modality *m* ∈ {1, …, *M*}:
  a. Draw a phenotype topic 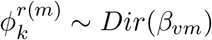 over the entire vocabulary *V* ^(*m*)^
2. For each EHR document *d* = {1, …, *D*}:
  a. Draw phenotype topic proportion from topic prior *θ*_*d*_ ∼ Dir(*α*_*d*_)
  b. For each EHR observation *i* = {1, …, *N*_*d*_} of any modality *m* ∈ {1, …, *M*}:
    i. Draw topic assignment *z*_*di*_ ∼ Mult(*θ*_*d*_)
    ii. Draw an EHR observation:

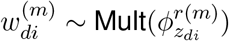

#### 10.2 MixEHR-SAGE methodology

##### 10.2.1 Initialization of phenotype topic priors

For each reference phenotype *k*, we run a two-component GMM on PheCode frequencies among patients. The normalized posterior probabilities of the higher components from the two components are then utilized as the initial topic prior 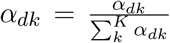 [60]. In most cases, Patients who have no recorded PheCode occurrence are unlikely to have the disease, thus the topic prior *α*_*dk*_ is set to zero for these patients. Based on the calculated topic priors, the sufficient statistics with regards to the ICD modality are initialized as follows:

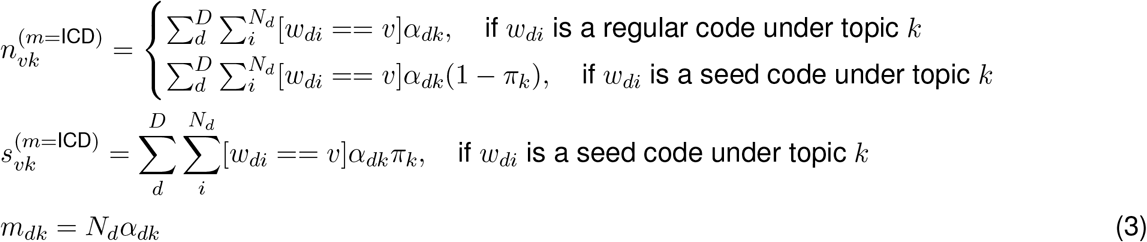

where *n*_*vk*_ denotes the number of times word *v* is assigned to a regular topic *k, s*_*vk*_ is the number of times the seed word *v* is assigned to seed topic *k*, and *m*_*dk*_ represents the estimated topic probabilities for each EHR document *d*.

For the unguided modalities, MixEHR-SAGE only needs to update the sufficient statistics 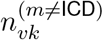:

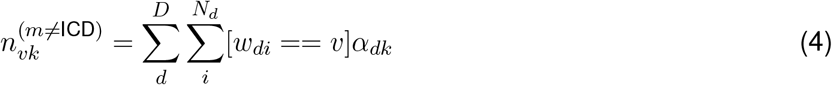

##### 10.2.2 Collapsed variational inference algorithm

In this section, we present the variational inference algorithm that approximates the posterior distribution of latent variables in the proposed MixEHR-SAGE. To reduce probabilistic dependencies among latent variables, we integrated out the latent variables *θ* by exploiting the conditional independency in the PGM (Figure 1) and the conjugate relationship between Dirichlet variables *θ* and multinomial topic assignment variables *z*. Similarly, we integrated out Dirichlet variables *ϕ*^*r*^ and *ϕ*^*s*^ due to its conjugacy to the multinomial EHR feature variables *w*. Note that we can always compute the sufficient statistics of these Dirichlet variables given the expectations of the multinomial variables and the hyperparameters (see Eq. (1)). For the *K*-dimensional hyperparameters *π*, we used empirical Bayes to optimize their fixed point estimates.

To infer the latent variables *z*, we applied variational inference by turning our inference problem into an optimization problem over an evidence lower bound (ELBO) of marginal likelihood:

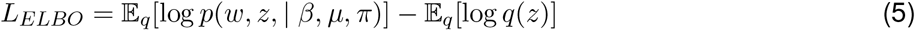

Under the independent mean-field assumption, we proposed a fully factorized variational family for the latent variables *z* and *η*:

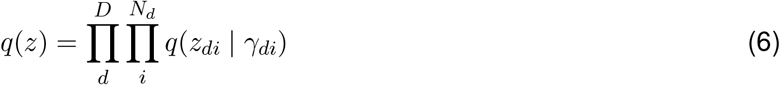

where the variational density of *z* is multinomial. Therefore, we employed mean-field, collapsed variational inference to optimize the variational parameters of *q*(*z*).

Regardless of the modality, the variational parameter of the topic assignment *γ*_*dik*_ for EHR feature *i* in EHR document *d* under phenotype topic *k* is defined as:

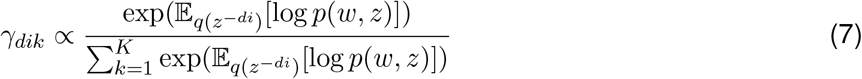

where the notation −*di* means the exclusion of token *i* in record *d*.

For the ICD modality, we take into account four scenarios when inferring the topic assignment of token *i* in document *d* under phenotype topic *k*:

1. If *w*_*di*_ is a seed ICD code under topic *k*, the posterior probability it is sampled from the *seed* topic distribution of topic *k* is:

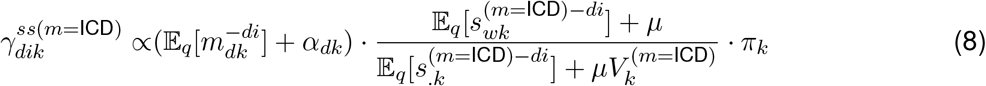
2. If *w*_*di*_ is a seed ICD code under topic *k*, the posterior probability it is sampled from the *regular* topic distribution of topic *k* is:

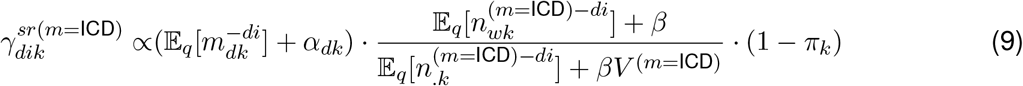
3. If *w*_*di*_ is a regular ICD code w.r.t. topic *k*, the posterior probability it is sampled from the *seed* topic distribution of topic *k* is 0.
4. If *w*_*di*_ is a regular ICD code w.r.t. topic *k*, the posterior probability it is sampled from the *regular* topic distribution of topic *k* is:

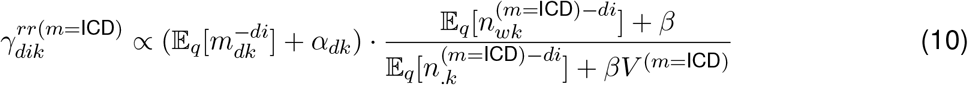

where *n*_.*k*_ and *s*_.*k*_ are simply the summation of the sufficient statistics *n*_*wk*_ and *s*_*wk*_ over all features. The expected values of sufficient statistics are computed as follows:

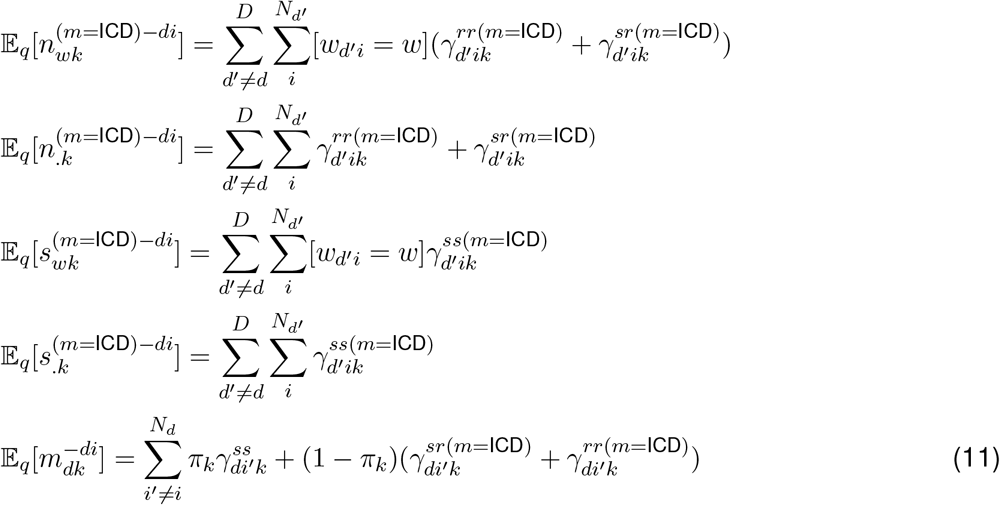

Note that the inference of regular topics by Eq. (10) benefits from guided information through 𝔼_*q*_[*m*_*dk*_], whose calculation relies on both seed topics and regular topics. This interdependency is the driving force behind our guided mechanism. Finally, the variational probabilities are normalized such that 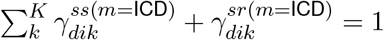 and 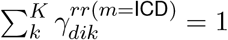:

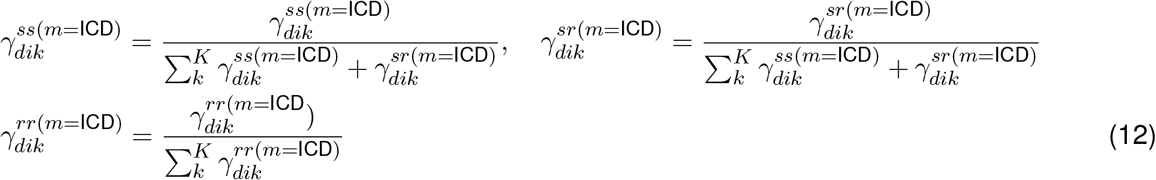

The seed-topic rates *π* are estimated by maximizing the marginal likelihood function under the variational expectations:

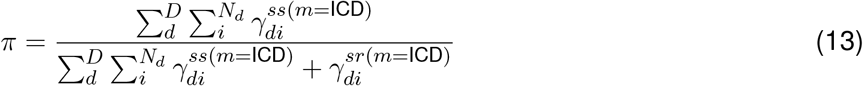

To infer latent topic assignments of other unguided modalities, the variational parameter 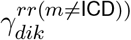 is updated without incorporating expert-guided knowledge:

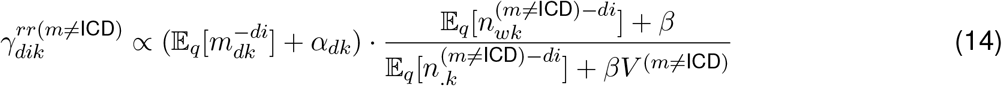

This variational update relies on *document-level* sufficient statistics 𝔼_*q*_[*m*_*dk*_] so that the topic inference for the unguided modalities also benefits from seed-guided information.

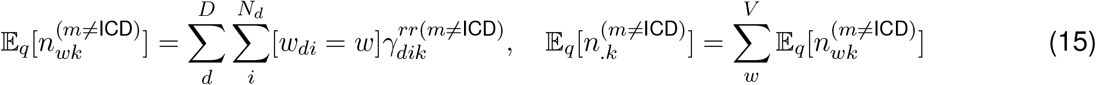

The complete inference algorithm is summarized in Algorithm 1. To effectively handle large-scale data, we performed stochastic variational inference using mini-batches containing 1,000 EHR documents per batch [48]. We used the validation set to fine-tune the topic hyperparameters *µ* and *β* to minimize the held-out negative log-likelihood. Upon convergence of the ELBO, we can compute the collapsed variables (*θ, ϕ*^*r*^, *ϕ*^*s*^) with the respective variational expectations:

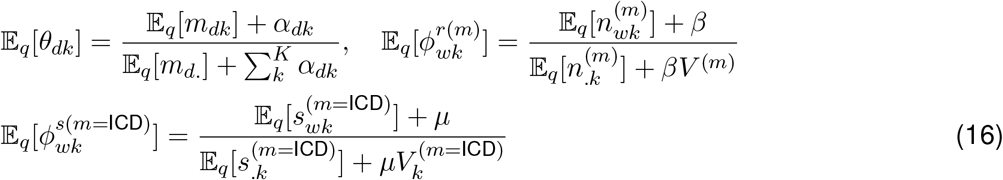

#### 10.3 MIMIC-III analysis

##### 10.3.1 MIMIC dataset

MIMIC-III is a clinical database that contains data from 38,597 adult patients and 7870 neonates admitted to the intensive care unit (ICU) of the Beth Israel Deaconess Medical Center between 2001 and 2012. The MIMIC-III dataset comprises a heterogeneous collection of EHR data such as diagnoses, laboratory test results, procedures, medications, and clinical notes for a total of 53,423 distinct hospital admissions [28]. The dataset can be downloaded from the [PhysioNet](http://mimic.physionet.org/) portal and used in accordance with the PhysioNet user agreement. This step allows for a rigorous assessment of MixEHR-SAGE’s ability to infer meaningful phenotype structures before its application to large-scale biobank data.

##### 10.3.2 Qualitative phenotyping evaluation in the MIMIC-III dataset

We qualitatively evaluated the inferred topic on the MIMIC-III dataset. We selected 9 common diseases for analysis: asthma (495), congestive heart failure (CHF, 428), chronic obstructive pulmonary disease (COPD, 496), diabetes (250), epilepsy (345), HIV (71), hypertension (401), ischemic heart disease (IHD, 411) and schizophrenia (295.1). For each phenotype, we extracted top 3 EHR features with highest probabilities across modalities (Figure S1A, C). We also compared the inferred disease topic from the MixEHR-Guided model to evaluate topic quality (Figure S1B, D).

We found that the inferred phenotype topics *ϕ*^*r*^ confer high probabilities to seed ICD codes for the selected phenotypes, indicating that expert-guided topic inference helps discover clinically meaningfully disease topics. For instance, the top ICD-9 codes for the schizophrenia topic were its seed ICD codes, 295.90, 295.70, and 295.30, matched the seed codes defined by its corresponding PheCode. The CHF topic assigned high probabilities to ICD codes 428.23, 428.22, 428.21, corresponding to complications of systolic heart failure. Another key finding was that MixEHR-SAGE assigned highest probabilities to non-seed ICD codes, revealing additional clinically relevant features. For example, the asthma topic assigned high probability to esophageal reflux, consistently with epidemiological and Mendelian randomization studies linking childhood-onset asthma to increased risk of gastroesophageal reflux disease [61, 62]. Similarly, the hypertension topic assigned high probability to the non-seed ICD code chronic kidney disease not otherwise specified, suggesting it commonly co-occurs with hypertension in ICU patients. We also found that high-probability ICD codes were generally consistent within the same disease systems. For example, 8 of the 9 top codes for CHF, hypertension and IHD, fell under the circulatory system category (Figure S1A).

Furthermore, MixEHR-SAGE inferred clinically meaningful phenotype topics for medication codes, even without using seed codes for this modality (Figure S1C). For example, the top drug codes for diabetes topics included insulin, commonly used to manage blood glucose in diabetic patients. We also identified a top medication prednisone, a corticosteroid used to decrease inflammation in asthma and COPD. Additional high-probability codes from other unguided modalities—-such as Current Procedural Terminology (CPT), Diagnosis Related Group (DRG), lab test, and doctor notes-—are shown in Figure S2.

We also compared the inferred topics from the baseline MixEHR-G method (Figure S1B, D). Most of the top ICD-9 codes were seed codes, while some frequently occurring non-seed ICD codes were incorrectly assigned high probabilities across unrelated disease topics. For instance, the non-seed ICD code 401.9 (Hypertension NOS) appeared with high probability in unrelated diseases like schizophrenia or epilepsy. This issue was more obvious for the medication modality, where unrelated drug codes like saline solutions were assigned high probabilities across nearly all disease topics, indicating the less specificity in the inferred topics from MixEHR-Guided (Figure S1D).

## Supplementary information

### S1 Supplementary Tables

**Table S1:**
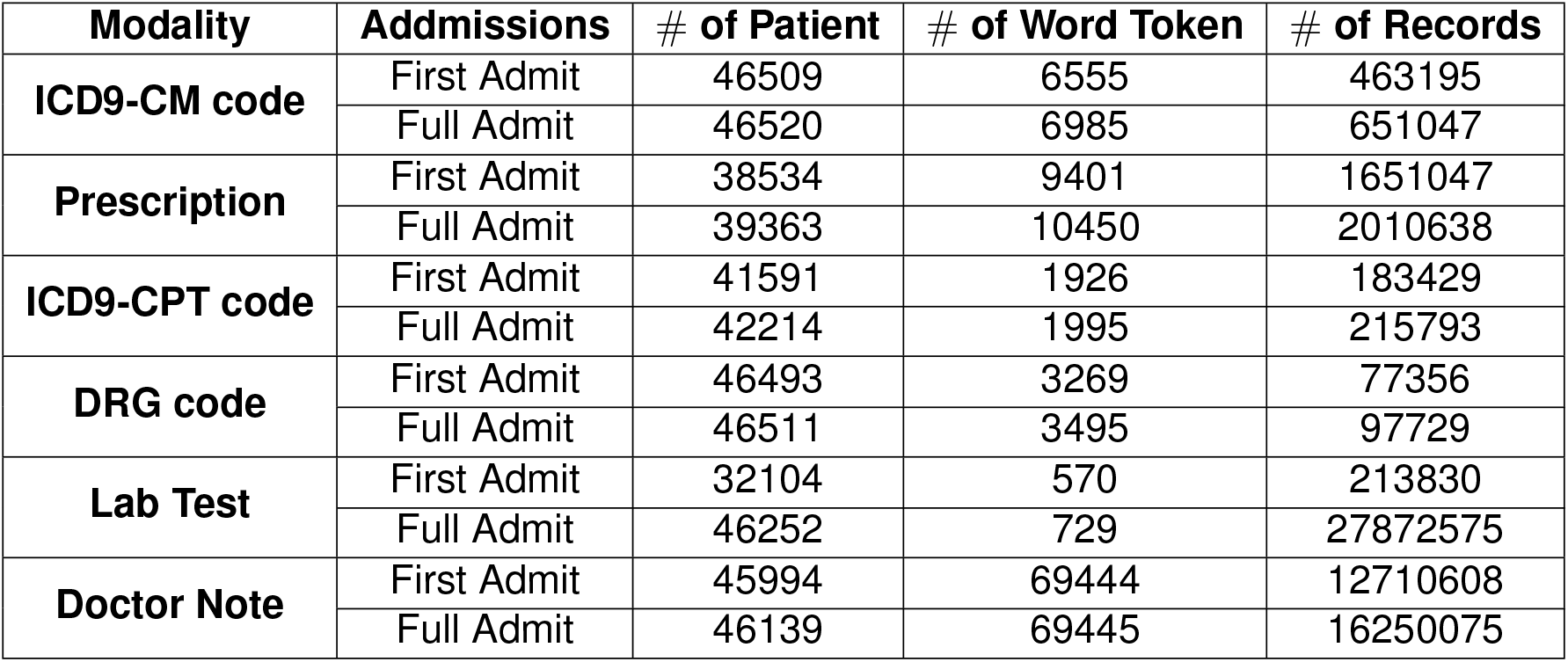
Summary of first and full admit data for processed MIMIC-III database

| Modality | Admissions | # of Patient | # of Word Token | # of Records |
| --- | --- | --- | --- | --- |
| ICD9-CM code | First Admit | 46509 | 6555 | 463195 |
|  | Full Admit | 46520 | 6985 | 651047 |
| Prescription | First Admit | 38534 | 9401 | 1651047 |
|  | Full Admit | 39363 | 10450 | 2010638 |
| ICD9-CPT code | First Admit | 41591 | 1926 | 183429 |
|  | Full Admit | 42214 | 1995 | 215793 |
| DRG code | First Admit | 46493 | 3269 | 77356 |
|  | Full Admit | 46511 | 3495 | 97729 |
| Lab Test | First Admit | 32104 | 570 | 213830 |
|  | Full Admit | 46252 | 729 | 27872575 |
| Doctor Note | First Admit | 45994 | 69444 | 12710608 |
|  | Full Admit | 46139 | 69445 | 16250075 |

**Table S2:** Notations in MixEHR-SAGE

| Notations | Descriptions |
| --- | --- |
| $D$ | total number of EHR documents in dataset |
| $N_d$ | total number of features for EHE document $d$ |
| $M$ | total number of modality in dataset |
| $K$ | number of phenotype topics |
| $V^m$ | feature vocabulary for modality $m$ in dataset |
| $V_k^{(m=ICD)}$ | set of seed ICD codes for phenotype topic $k$ |
| $w_{di}$ | feature index of token $i$ in EHR document $d$ |
| $z_{di}$ | topic assignment for feature $w_{di}^{(m)}$ |
| $x_{di}$ | binary seed-topic indicator of ICD code $w_{di}^{(m=ICD)}$ |
| $\alpha \in R^{D \times K}$ | Dirichlet topic priors |
| $\theta \in R^{D \times K}$ | topic mixture memberships |
| $\phi^r \in R^{K \times V}$ | regular topic distributions |
| $\phi^s \in R^{K \times S}$ | seed topic distributions |
| $\pi \in R^K$ | seed-topic rates |
| $\eta$ | topic hyperparameter for regular topics $\phi^r$ |
| $\mu$ | topic hyperparameter for seed topics $\phi^s$ |

Table S3: Genome-wide significant loci identified by MixEHR-SAGE versus traditional binary GWAS approach for the top 100 most prevalent phenotypes. The table was provided as a CSV file named “MixEHR-SAGE_vs_Binary_Loci_Comparison” in the supplementary materials.

### S2 Supplementary Figures

**Figure S1:**
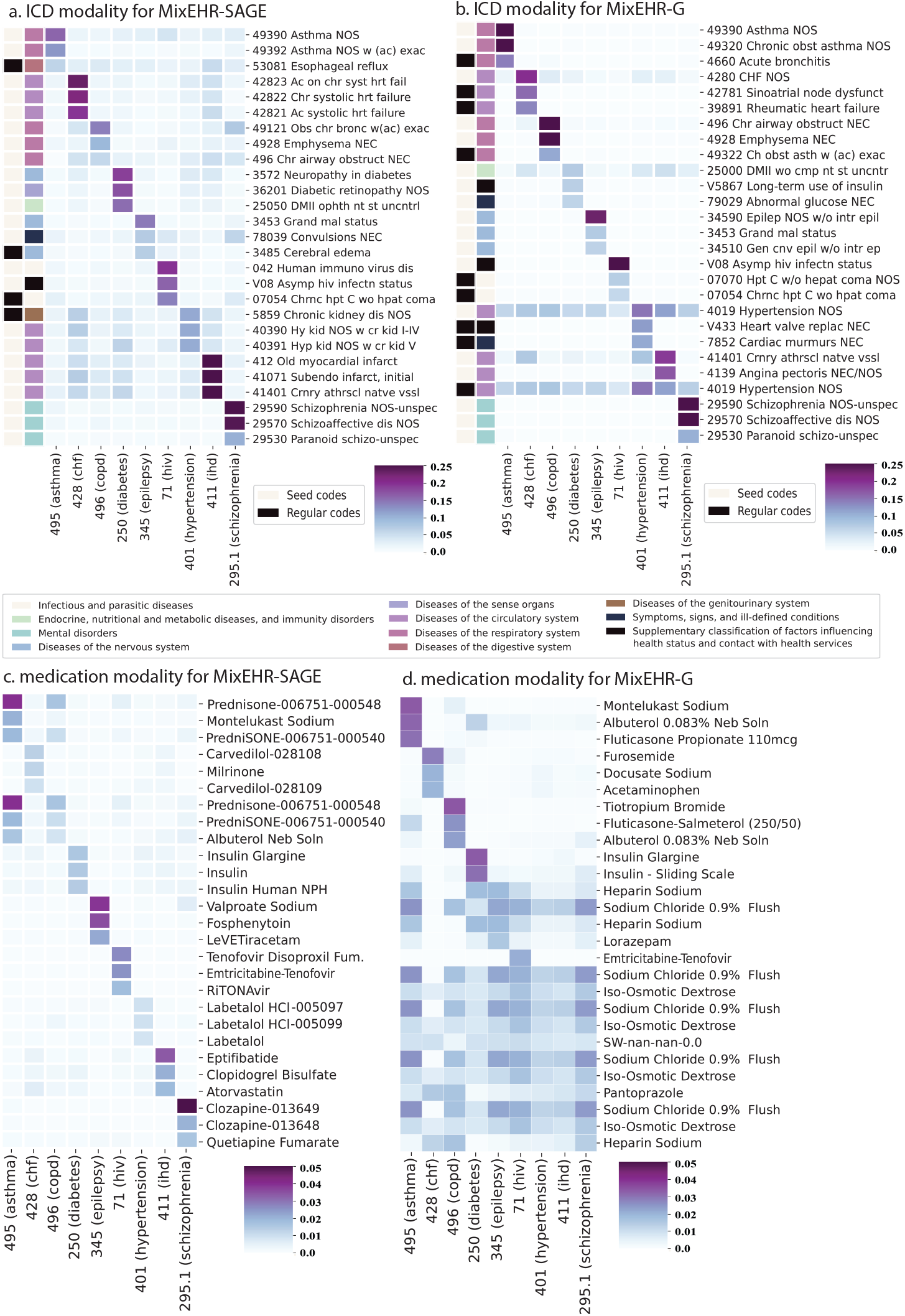
The 3 highest probability ICD and drug codes for topics inferred by MixEHR-SAGE and MixEHR-Guided on MIMIC-III data. The top ICD diagnostic codes are presented for (a) MixEHR-SAGE and (b) MixEHR-Guided. The colorbars represent different ICD categories and whether an ICD code is a seed or regular code. The top 3 drugs for the 9 selected phenotype topics as identified by (c) MixEHR-SAGE and (d) MixEHR-Guided

**Figure S2:**
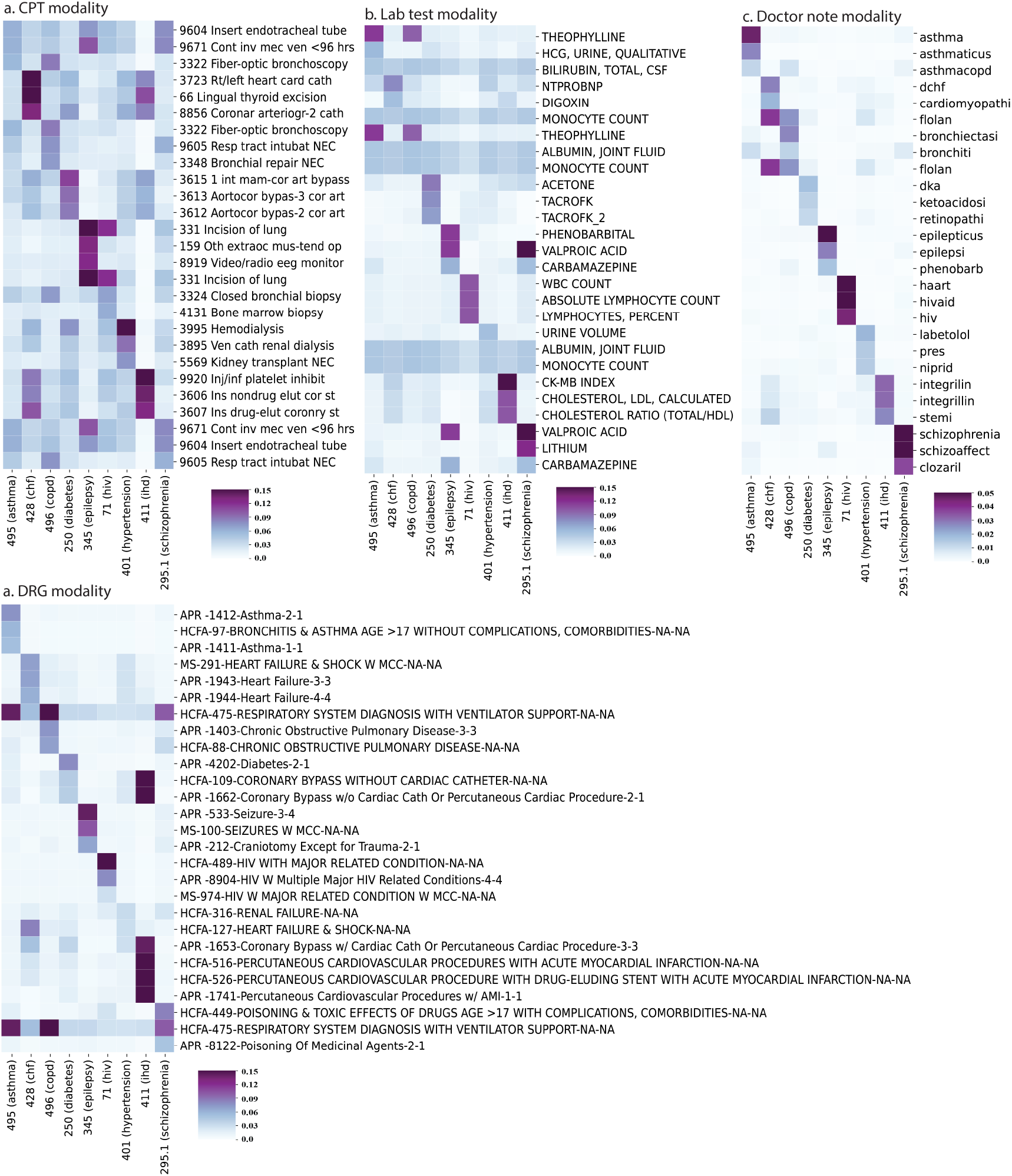
The 3 highest probability EHR codes (CPT, DRG, lab tests and doctor notes) for topics inferred by MixEHR-SAGE on MIMIC-III data.

**Figure S3:**
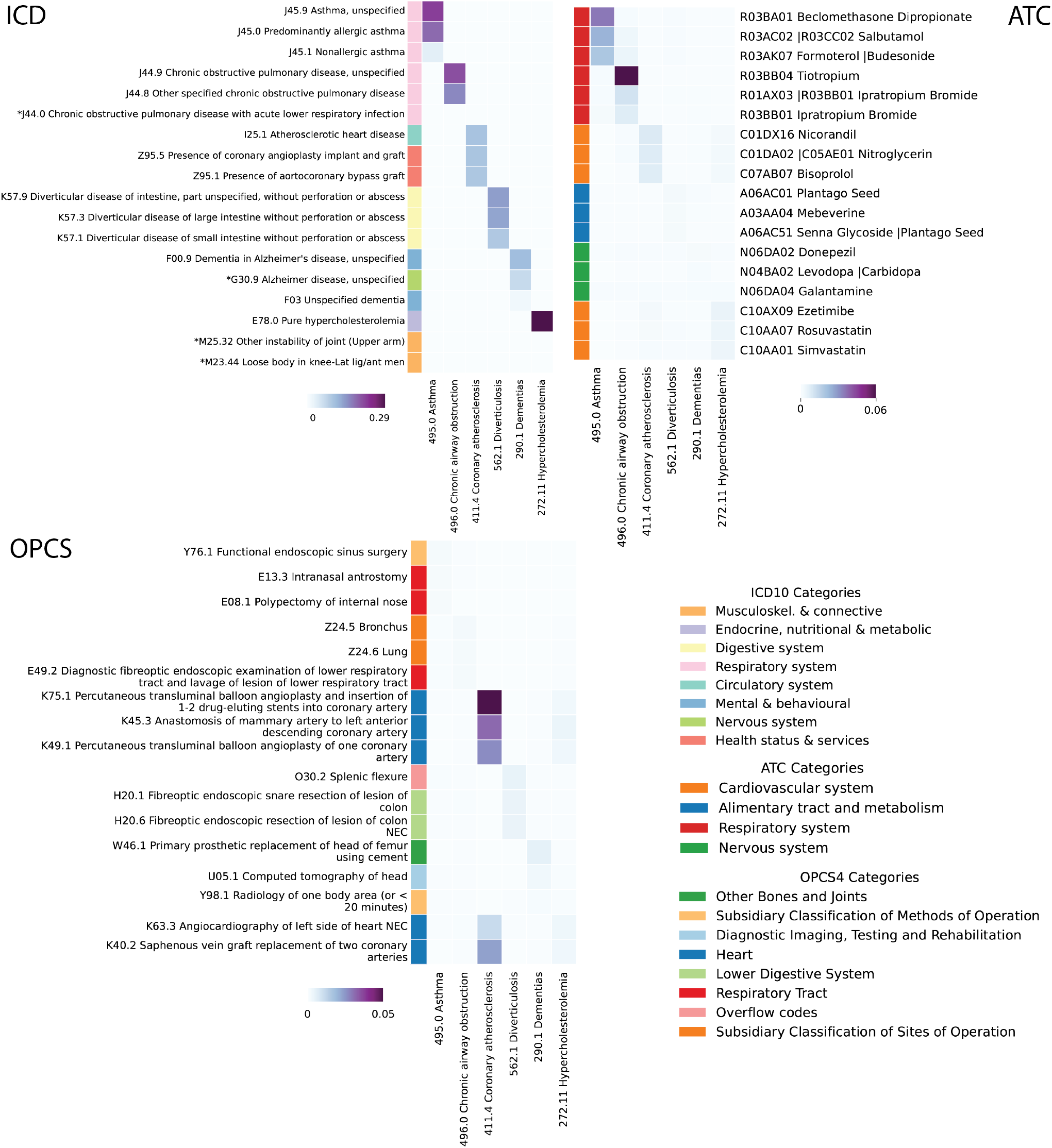
The 3 highest probability EHR codes (ICD, ATC, OPCS4 code) for different topics identified by MixEHR-SAGE on UKB data.

**Figure S4:**
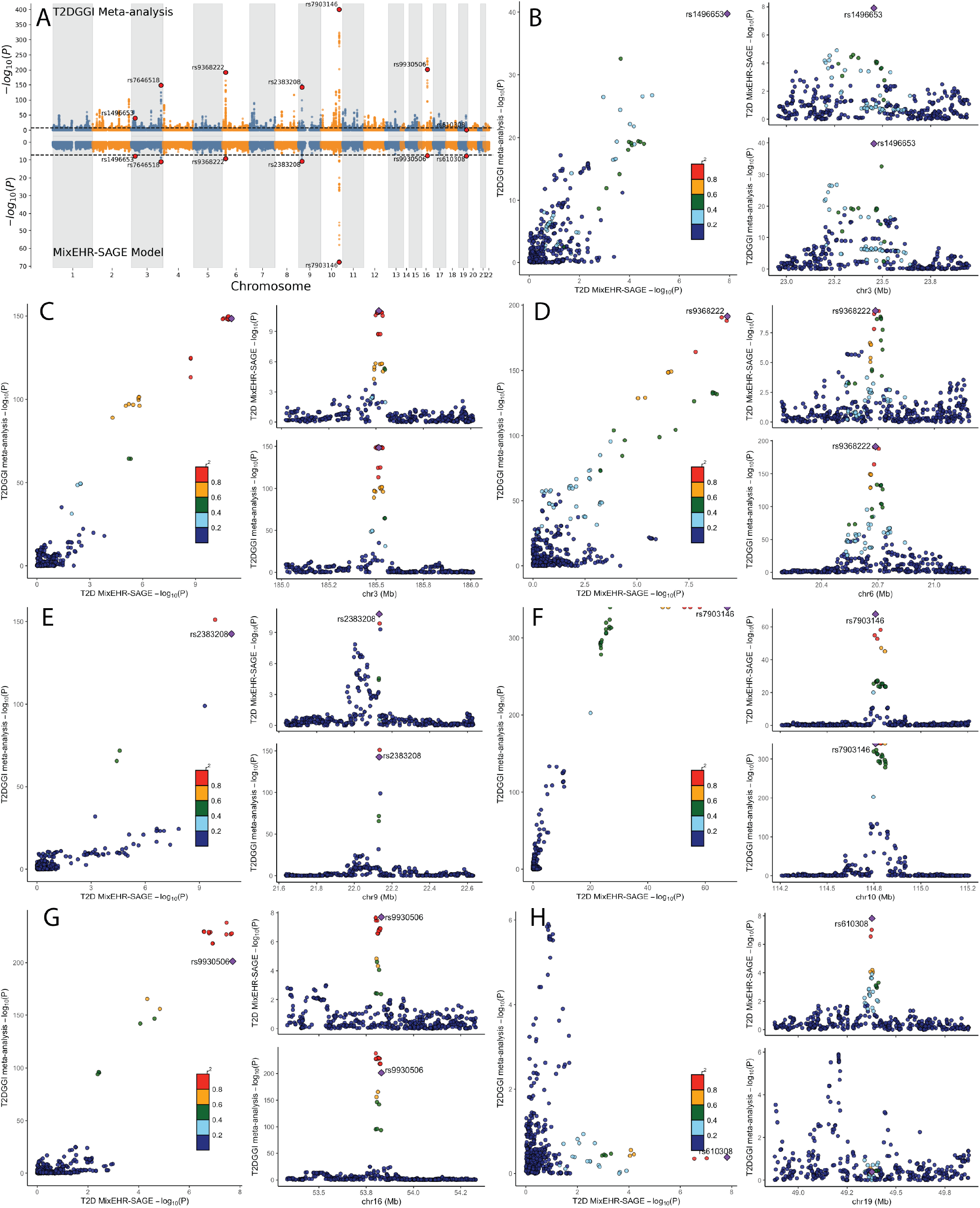
Genome-wide and locus-specific comparison of T2D associations between the T2DGGI meta-analysis and the MixEHR-SAGE model. (A) Miami plot displaying GWAS comparison between T2DGGI meta-analysis and MixEHR-SAGE. Genome-wide significant loci were highlighted with their lead SNPs in red. (B–H) Regional plots for the genome-wide significant loci highlighted in panel A. For each subpanel, the left shows the locuscompare scatter plots of −log_10_(*P*) values between the two studies with LD coloring (*r*^2^ from 1000 Genomes EUR reference panel). The right subpanel shows regional association plots centered on the lead SNP.

## Notes

### Competing Interest Statement

The authors have declared no competing interest.

### Summary of Updates

Improved figures. Validation results on external GWAS

